# Activating variants in PRKD2 in two families with Systemic sclerosis, and enrichment of rare variants in a sporadic series

**DOI:** 10.64898/2026.09.20.26363195

**Authors:** Pierre Maus, Gaëlle Tilman, Simon Boutry, Axelle Loriot, Thomas Conjard, Jinine Hatoum, Anne-Catherine Dens, Tessa Du Four, Delphine Nolf, Thibault Hirsch, Tiphanie Gomard, Raphaël Helaers, Ellen De Langhe, Yuki Ishikawa, Yannick Allanore, Chikashi Terao, Jacques Fellay, Marie Vanthuyne, Vanessa Smith, Nisha Limaye

## Abstract

Systemic sclerosis (SSc) is a rare, heterogeneous autoimmune disease characterized by immune dysregulation, vasculopathy and fibrosis. While typically sporadic, SSc can in a minority of cases show familial clustering. Whole exome sequencing (WES), performed on six families with two affected members each, identified a total of 253 genes with rare, disease co-segregating variants predicted to affect protein function. Of these, we prioritized *PRKD2*, one of only three genes identified in two families. *PRKD2* encodes Protein kinase D2: an intracellular serine-threonine kinase that amplifies T cell receptor (TCR) signaling. Both familial SSc-associated PRKD2 missense variants increased kinase activation, exerting overlapping, activating effects on TCR signaling in heterozygous knock-in Jurkat cells. A pan- PRKD inhibitor suppressed T cell activation in peripheral blood mononuclear cells from both familial SSc cases and controls. The enrichment of *PRKD2* rare variants in a sporadic SSc cohort provides further support for its relevance to SSc pathogenesis. Together, these findings identify *PRKD2* rare variants as potential genetic contributors to SSc, and PRKD2-mediated TCR signaling as a candidate disease-relevant and potentially targetable pathway.

## Introduction

Systemic sclerosis (SSc) is an autoimmune disease characterized by small vessel vasculopathy, immune dysregulation, and fibrosis of the skin and internal organs^1^. While relatively rare (prevalence 4-34 per 100,000 individuals^2, 3^) and phenotypically heterogeneous, SSc is associated with significant morbidity, heavy disease burden, and the highest mortality rate amongst rheumatic diseases^3^. Criteria for early diagnosis include objective Raynaud’s phenomenon with characteristic vascular changes in the digits (scleroderma-pattern) detected by nailfold capillaroscopy, and/or the presence of SSc-specific autoantibodies^1^. About 70% of patients who fulfill the criteria for early, limited SSc (lSSc) develop “definite” SSc within a five to ten year period^4^, sub-classified according to the extent of skin involvement. In limited cutaneous SSc (lcSSc), skin fibrosis is limited to the distal members; in diffuse cutaneous SSc (dcSSc), skin fibrosis is extensive and progressive. These forms also tend to differ in disease course: time to develop first non-Raynaud’s symptoms, rate of progression, types of autoantibodies, internal organ involvement, and causes and risk of mortality^1^.

Current therapies for SSc are largely aimed at symptom control, with sparse evidence for *bona fide* disease-modifying effects. Standard immunosuppressive therapies provide limited benefit, with autologous hematopoietic stem cell transplantation, used in only the most severe cases, being the only therapy shown to improve survival^5^. More recently, the use of CD19 (B cell)- targeting chimeric antigen receptor (CAR) T-cells has shown early promise, including amelioration of skin and organ fibrosis^6^. This supports a central role for adaptive immunity in the pathogenesis of SSc, as do data from large-scale genome wide association studies (GWAS): The HLA class II region shows the strongest associations with disease phenotypes; the majority of non-HLA susceptibility loci also map to genes with diverse immune functions^7–9^.

Deciphering how particular susceptibility alleles contribute to disease, and how this may usefully be leveraged in disease-modifying therapies for (subsets of) patients, remains challenging.

While typically sporadic, SSc can in rare cases show familial clustering; genetic analysis of such families has the potential to identify high-impact disease-cosegregating variants, with biological effects amenable to functional testing in cellular or animal models. We performed whole exome sequencing (WES) on blood-DNA from the members of six duplex SSc families, from two large Belgian centers of expertise (Ghent University Hospital and Cliniques universitaires St Luc). Here, we report the identification and functional testing of heterozygous missense rare-variants in the *PRKD2* gene, identified in two of the six families.

*PRKD2* encodes Protein Kinase D2: one of three intracellular serine/threonine kinases of the Protein Kinase D family that are activated by diacylglycerol (DAG) and Protein Kinase C (PKC), in myriad cell signaling contexts^10, 11^. PRKD2 has been shown, in mouse models and human cell lines, to act as an amplifier of T cell receptor (TCR) signaling and T cell activation^11–15^. We investigated the functional effects of the two variants identified in familial SSc, in engineered (heterozygous knock-in) Jurkat CD4 T cells. Intriguingly, both variants increased PRKD2 kinase activation, with overlapping but not identical effects on TCR-signaling and T cell activation. WES performed on a sporadic SSc series identified an additional ten non-synonymous rare variants in the *PRKD2* gene, six of which were significantly enriched in cases as compared to gnomAD population controls. The gene-level rare variant enrichment of *PRKD2* in the series suggests that this axis may also be relevant in sporadic disease.

## Results

### Whole exome sequencing on six SSc families

The genetic analysis of familial forms of SSc is rendered challenging, in part, due to its rarity (<2% of cases in a relatively rare rheumatic condition^16^). We performed WES on blood DNA from twelve affected and two unaffected members of six families (**Fig.1A**). The series shows considerable intra- and inter-familial phenotypic heterogeneity (**Fig.1A-B**; a more detailed clinical description is provided in **Supplementary Table S1**). Each family includes two affected first-degree relatives; this is in keeping with previously reported families (**Supplementary Table S2**), the majority of which are also duplex. Phenotypic heterogeneity, together with a paucity of large and/or multi-generational pedigrees in familial SSc, may suggest di- or oligogenic inheritance: two or more randomly segregating genes mediating disease, or modifying disease expressivity mediated by a primary “driver” ^17^.

**Figure 1.**
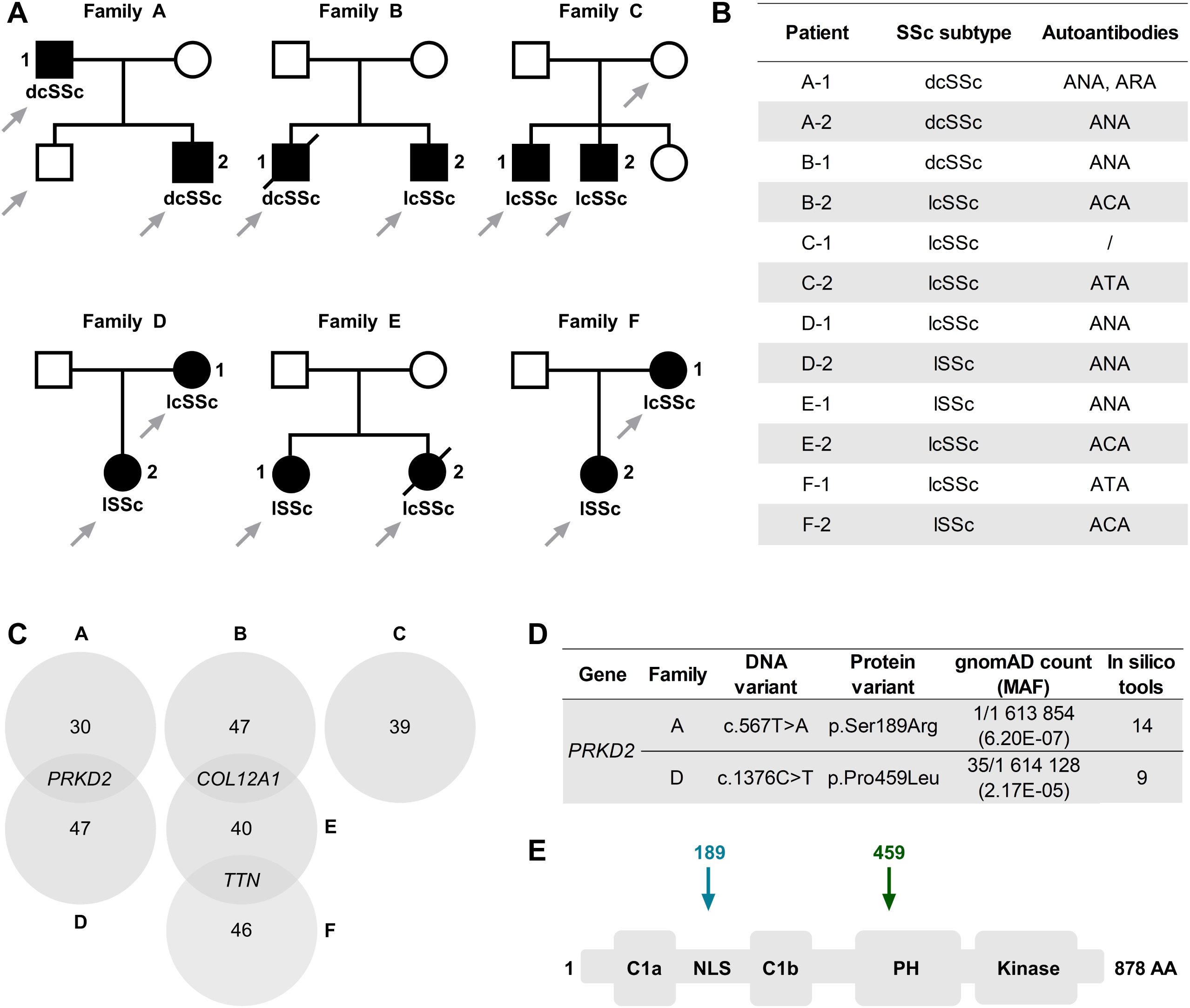
WES performed on six SSc families identifies *PRKD2* as a candidate gene in two. (**A**) Six SSc families. Filled symbols: affected. Clear symbols: unaffected. Strikethrough: deceased. Arrows: WES performed on blood-DNA. **lSSc:** limited SSc. **lcSSc:** limited cutaneous SSc. **dcSSc:** diffuse cutaneous SSc. (**B**) Summary of clinical phenotypes (detailed description in **Supplementary Table S1**). ANA: anti-nuclear antibodies, ACA: anti-centromere antibodies, ATA: anti-topoisomerase I antibodies, ARA: anti-RNA polymerase III antibodies. (**C**) Venn diagram showing the number of candidate genes retained per family by WES variant filtering. (**D**) *PRKD2* variants identified in (both affected members of) Families A and D. The variant in Family A is also shared by the unaffected member for whom WES was performed. gnomAD count: number of variant alleles/total number of genotyped alleles (*MAF*: Minor Allele Frequency) in v4.1.0. Both variants are absent in all other samples in the local WES database. *In silico* tools: Number of programs (out of 20) that predict missense variants are “pathogenic” to protein function. (**E**) Schematic representation of PRKD2 protein domains indicating the location of the amino acid substitutions identified in Family A (189, blue) and D (459, green). AA: amino acid.

With this in mind, WES data were filtered to retain all variants *(i)* shared by both affected members within each family, regardless of whether they are also shared by any available unaffected members, *(ii)* with minor allele frequency (*MAF*) ≤ 0.001 in the general population (public databases and the in-house WES database of the Genomics Platform of UCLouvain), *(iii)* predicted to affect protein function (nonsense and consensus splice-site variants, missense variants predicted “pathogenic” by five or more *in silico* tools) (**Supplementary Table S3**). This yielded a total of 256 variants in 253 genes across the six families (31-48 variants per family, **Fig.1C**, **Supplementary Table S4**). Only three genes: *TTN*, *COL12A1* and *PRKD2*, were “shared”, with variants satisfying these criteria identified in two families each (**Supplementary Table S5**). *TTN* is the largest protein-coding gene in the human genome (cDNA>100kb), with the highest rate of non-synonymous variants in the general population^18^.

As a result, it is frequently identified by typical WES variant-filtering strategies, across phenotypes (observation in our lab and others). Variants in *TTN* (MIM#188840) have been linked to a range of (cardio)myopathies. *COL12A1*, which codes for a large fibril-associated collagen, has also been linked to certain rare congenital myopathies (MIM#616471 and 616470). As an ECM component, it is a potentially interesting candidate mainly for fibrosis, observed in only two of the four individuals with variants in this gene (**Fig.1B**, **Supplementary Table S1**). Given its role in adaptive immunity, dysregulation of which is common to all SSc sub-types, we instead prioritized the *PRKD2* gene for further functional analysis. We began with full-length PCR-amplification and sequencing of *PRKD2* cDNA from PBMC from the patients in both families. This confirmed the expression of both alleles, with no apparent splice- effects exerted by either variant (**Supplementary Figure S1**).

### p.Ser189Arg and p.Pro459Leu show similar effects on PRKD2 auto-phosphorylation but distinct effects on nuclear translocation in heterozygous knock-in Jurkats

The PRKD2 protein consists of an N-terminal regulatory region (composed of DAG-binding domains, and a plekstrin homology (PH) domain that negatively regulates kinase activity), and a C-terminal kinase domain^19–22^. The variant identified in Family A (c.567T>A, p.Ser189Arg; NM_016457) is located in a nuclear localization signal (NLS), between the C1a and C1b DAG-binding regions^22, 23^ (**Fig.1D,E**). The variant in Family D (c.1376C>T, p.Pro459Leu) is located in the PH domain (**Fig.1D,E**). Both are highly conserved across species (not shown). Activation of PRKD2 requires plasma membrane recruitment by DAG, produced by PLCγ downstream of TCR-stimulation^20–22^ (**Supplementary Figure S2**). There, PRKD2 undergoes auto-transphosphorylation on Ser876, with phosphorylation of Ser706 and Ser710 by PKC required for full activation^10, 22, 24, 25^. Activated PRKD2 feeds into (but is not required for) the activation of the NFAT and NF-κB arms of TCR signaling, via unknown mechanisms^12, 13, 26–30^ (Supplementary Figure S2).

To test for the effects of the PRKD2 variants identified in familial SSc (p.Ser189Arg and p.Pro459Leu), we generated heterozygous knock-in (KI) Jukat cell-lines for each variant using CRISPR/Cas9-homology directed repair (HDR) followed by Next Generation Sequencing (NGS)-based screening. *PRKD2*-knockout (KO), non-targeted wild-type (WT) and scrambled guide-RNA (scRNA) nucleofected clones were generated as controls (3 clones per genotype). PCR-amplification and sequencing of full-length *PRKD2* cDNA confirmed the expression of both alleles in KI clones, with no aberrant splice-changes detected (not shown). Total levels of PRKD2 protein, assessed by Western blot, were similar between KI, WT and scRNA Jurkat cell lines; no expression was detected in KO lines (shown for one set of clones in **Fig.2A**).

**Figure 2.**
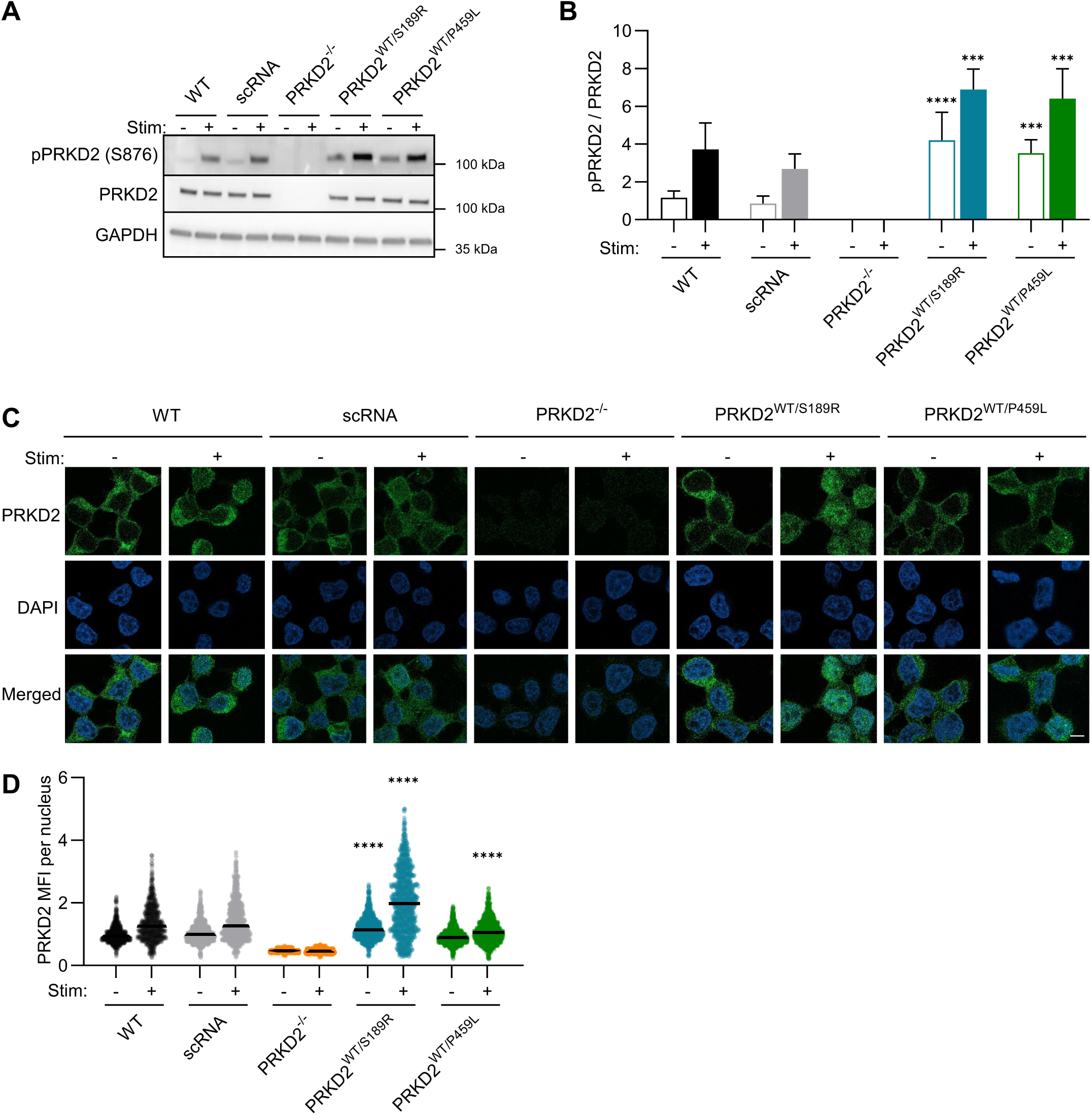
p.Ser189Arg and p.Pro459Leu increase protein phosphorylation and alter nuclear localization of PRKD2 in KI Jurkats. Jurkat CD4 T cell clones were serum-starved for 24hours in X-VIVO-10 media, then stimulated (+) or not (-) for 30 minutes with anti-CD3 (1µg/mL), anti-CD28 (1µg/mL) and the crosslinker STAR117 (5µg/mL). (**A-B**) Cells were lysed and Western blot performed for the indicated proteins. (**A**) Representative images and (**B**) Signal quantitation. pPRKD2/total PRKD2, each normalized to GAPDH performed on the same blot. n=3 experiments, each performed on 3 clones per genotype. \*\*\**p<0.001*, \*\*\*\**p<0.0001 vs.* corresponding WT, nested one-way ANOVA with Dunnett’s post-hoc multiple comparisons test. (**C-D**) Immunofluorescence staining for PRKD2 (green) and DAPI for nuclei (blue). (**C**) Representative images. (**D**) Mean Fluorescence Intensity (MFI) of PRKD2 staining per nucleus. Each dot represents one nucleus, black lines indicate the mean across all nuclei per genotype per condition. \*\*\**p<0.001*, \*\*\*\**p<0.0001 vs.* corresponding WT, two-way ANOVA with Dunnett’s post-hoc multiple comparisons test.

Both variants significantly increased basal activation of PRKD2 in (PRKD2^WT/S189R^ and PRKD2^WT/P459L^) heterozygous KI Jurkats, based on Western blot for the auto-phosphorylation site phospho-Ser876 (**Fig.2A-B**). A further increase of ∼1.5x was observed upon TCR stimulation of KI Jurkats. While phospho-PRKD2 levels remained lower than in KI Jurkats, WT and scRNA cells showed greater responsiveness to TCR-stimulation, with a 3-4x increase in phospho-PRKD2 as compared to their basal levels. This was not associated with differences in phosphorylation of the TCR-proximal signaling adaptors ZAP70 and LAT (**Supplementary Figure S3)**. The TCR-induced increase in PRKD2 auto-phosphorylation occurred earlier (by 5 minutes) in WT, scRNA and PRKD2^WT/S189R^ cells, and later (at 30 minutes of stimulation) in PRKD2^WT/P459L^ cells (**Supplementary Figure S4**).

TCR-stimulation results in nuclear translocation of PRKD2, from the cytoplasm where it is predominantly located at baseline^22, 23^. This was significantly increased in the case of PRKD2^WT/S189R^ cells, but not PRKD2^WT/P459L^ cells, which instead showed reduced nuclear translocation of PRKD2 as compared to their WT counterparts (**Fig.2C-D**). This may suggest that increased PRKD2 auto-phosphorylation, observed with both variants, does not suffice to modulate protein localization.

In summary, while both variants seem to enhance PRKD2 phosphorylation in heterozygous KI Jurkats, they exert distinct effects on its nuclear translocation.

### p.Ser189Arg and p.Pro459Leu alter TCR-induced activation of NFAT, NF-κB and ERK in heterozygous KI Jurkats

TCR-stimulation results in the activation of multiple pathways, with the amplitude, duration and relative strength of the NFAT and NF-κB arms described to play a major role in determining the nature of T cell responses elicited^31^. Ca^2+^ flux, a key step in NFAT activation, is significantly increased in PRKD2^WT/P459L^ cells as compared to all other cell lines (**Fig.3A– C**). For a more direct read-out of NFAT activation, we transduced all cell-lines with a lentiviral reporter construct for NFAT-inducible GFP and constitutive mCherry expression. In keeping with Ca^2+^ flux data, PRKD2^WT/P459L^ (but not PRKD2^WT/S189R^) cells showed significantly higher mCherry-normalized GFP expression, assessed by flow cytometry; PRKD2-deficient cells displayed reduced NFAT activation (**Fig.3D-E**). TCR-induced NF-κB activation, assessed by Western blot (for phosphorylation and degradation of IκBα and phosphorylation of NF-κB), was significantly increased by both variants in heterozygous KI as compared to control cells (**Fig.4A-D**). This was confirmed by flow cytometry on cells transduced with a lentiviral NF-κB reporter construct (**Fig.4E,F**). PRKD2-deficient cells showed decreased TCR-induced NF-κB activity.

**Figure 3.**
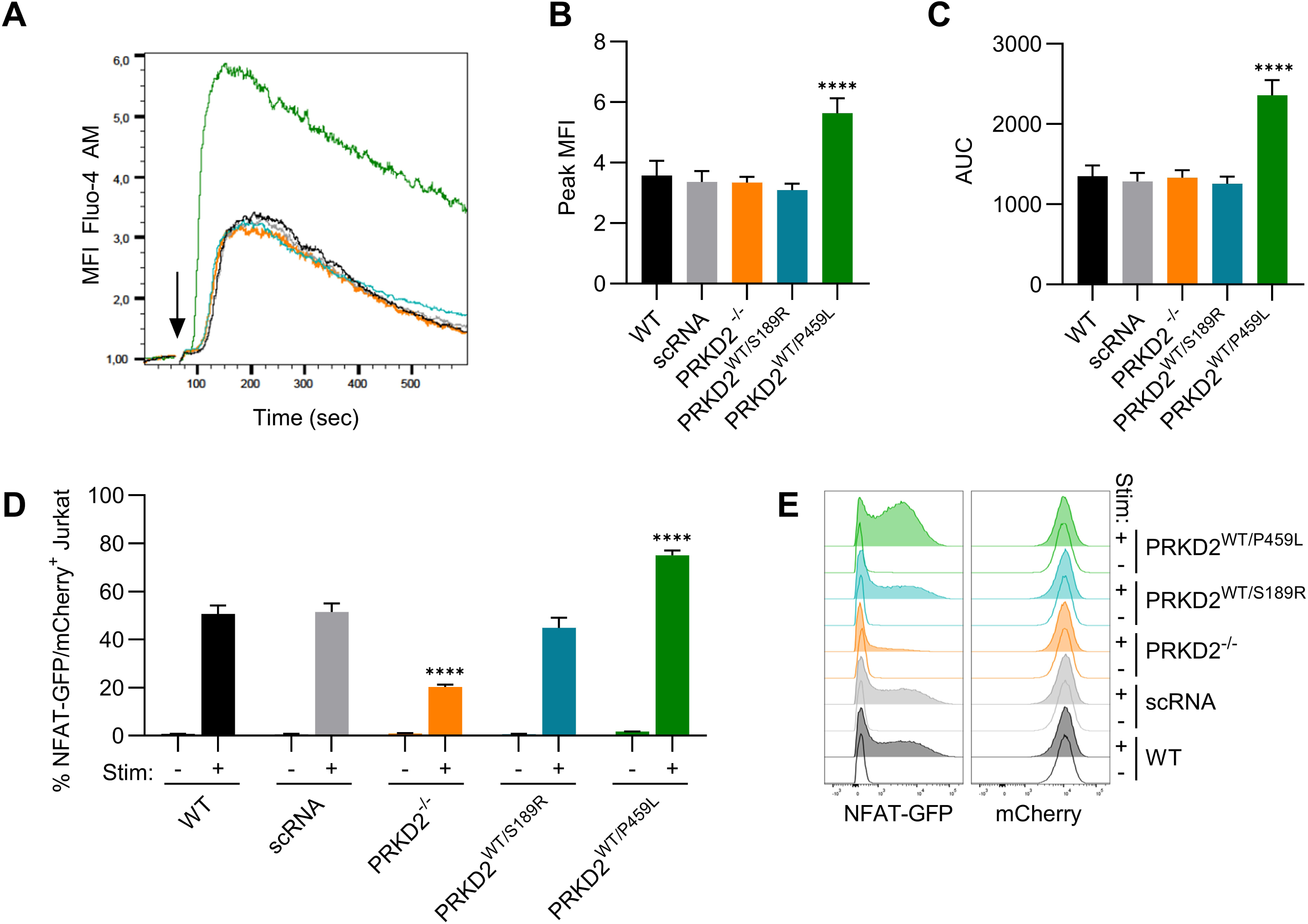
PRKD2 p.Pro459Leu, but not p.Ser189Arg, increases NFAT activation in TCR- stimulated KI Jurkats. (**A-C**) Serum-starved Jurkat CD4 T cell clones were incubated with the cytoplasmic calcium dye Fluo-4 AM. Fluorescence was measured by flow cytometry for 1 minute at baseline, and from 1 to 10 minutes following TCR stimulation (arrow) using anti- CD3 (1µg/mL), anti-CD28 (1µg/mL) and the crosslinker STAR117 (5µg/mL). (**A**) Representative fluorescence intensity data from one WT (black), scRNA (grey), PRKD2^-/-^ (orange), PRKD2^WT/S189R^ (blue), and PRKD2^WT/P459L^ (green) Jurkat clone each. (**B**) Peak MFIs and (**C**) AUCs (area under the curve) from n=3 experiments, each performed on 3 clones per genotype. (**D, E**) Clones were transduced with a vector encoding GFP (Green Fluorescent Protein) under an NFAT-responsive promoter and mCherry under a constitutive (PGK) promoter. GFP and mCherry expression levels were measured by flow cytometry after 16 hours of TCR stimulation as above. (**D**) Percentages and (**E**) representative histograms from n=3 experiments, each performed on 3 clones per genotype. \*\*\*\**p<0.0001* vs. corresponding WT, nested one-way ANOVA with Dunnett’s post-hoc multiple comparisons test.

**Figure 4.**
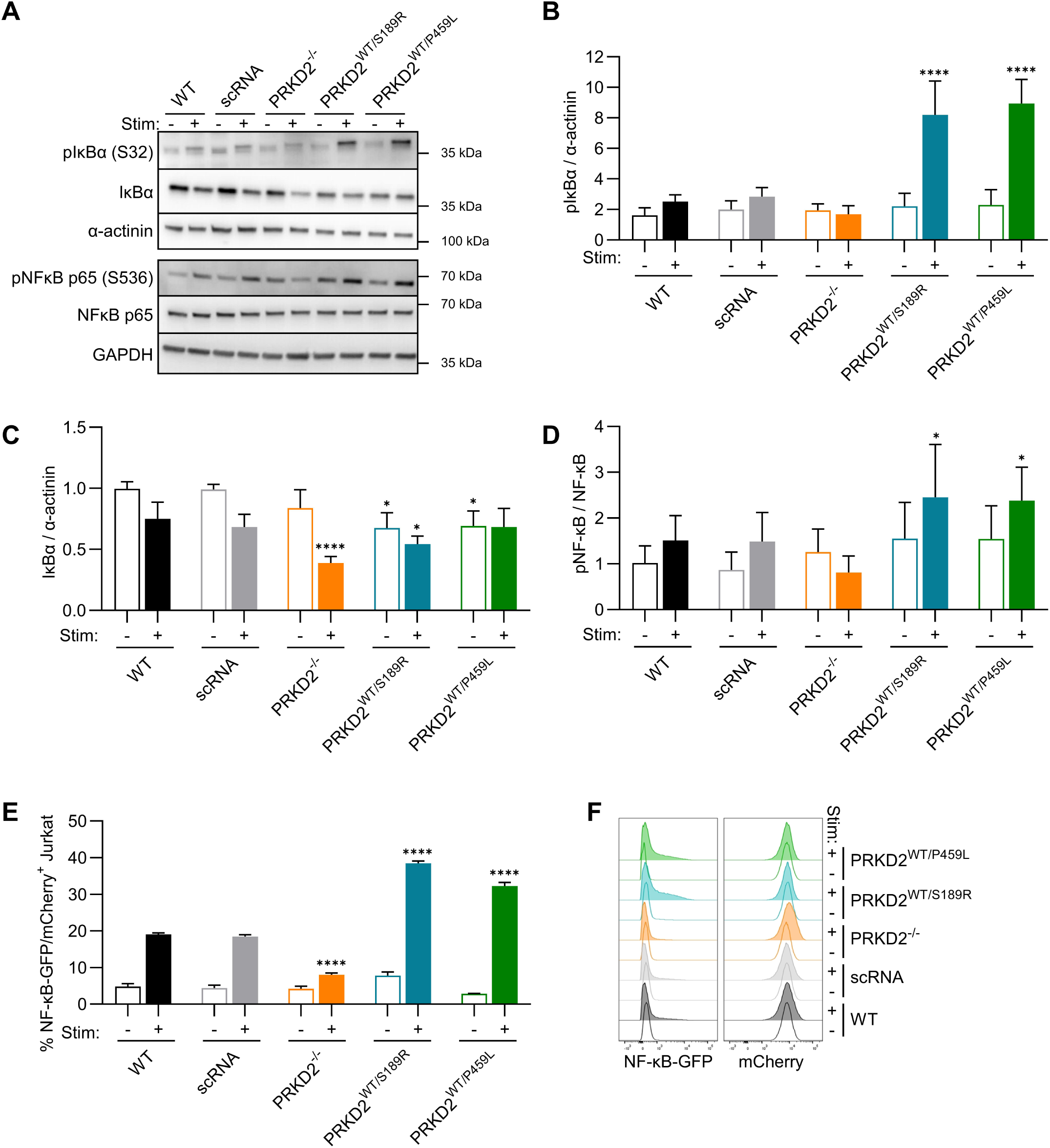
PRKD2 p.Ser189Arg and p.Pro459Leu increase NF-κB activation in TCR- stimulated KI Jurkats. Jurkat CD4 T cell clones were serum-starved for 24hours in X-VIVO- 10 media, then stimulated (+) or not (-) for 30 minutes with anti-CD3 (1µg/mL), anti-CD28 (1µg/mL) and the crosslinker STAR117 (5µg/mL). (**A-D**) Cells were lysed and Western blot performed for the indicated proteins. (**A**) Representative images and (**B-D**) Signal quantitation. (**B**) pIκBα and (**C**) total IκBα, normalized to α-actinin performed on the same blots. (**D**) pNF-κB/total NF-κB, each normalized to GAPDH performed on the same blot. n=3 experiments, each performed on 3 clones per genotype. \**p<0.05*, \*\*\*\**p<0.0001 vs.* corresponding WT, nested one-way ANOVA with Dunnett’s post-hoc multiple comparisons test. (**E-F**) Clones were transduced with a vector encoding GFP (Green Fluorescent Protein) under an NF-κB- responsive promoter and mCherry under a constitutive (PGK) promoter. GFP and mCherry expression levels were measured by flow cytometry after 16 hours of TCR stimulation as above. (**E**) Percentages and (**F**) representative histograms from n=3 experiments, each performed on 3 clones per genotype. \*\*\*\**p<0.0001 vs.* corresponding WT, nested one-way ANOVA with Dunnett’s post-hoc multiple comparisons test.

We next assessed the impact of PRKD2 variants on basal and TCR-induced ERK and AKT phosphorylation^32, 33^ (**Fig.5A-C**). A robust increase in ERK (p42/44) phosphorylation was observed upon TCR-stimulation of all cell lines, with PRKD2-deficient cells showing a significant decrease, and both PRKD2^WT/S189R^ and PRKD2^WT/P459L^ cells showing a significant increase as compared to controls (**Fig.5A,B**). The cell-lines showed uniformly high basal phosphorylation of AKT, perhaps due to a reported PTEN deficiency in Jurkats^34^, with no change observed upon TCR stimulation (**Fig.5A,C**).

**Figure 5.**
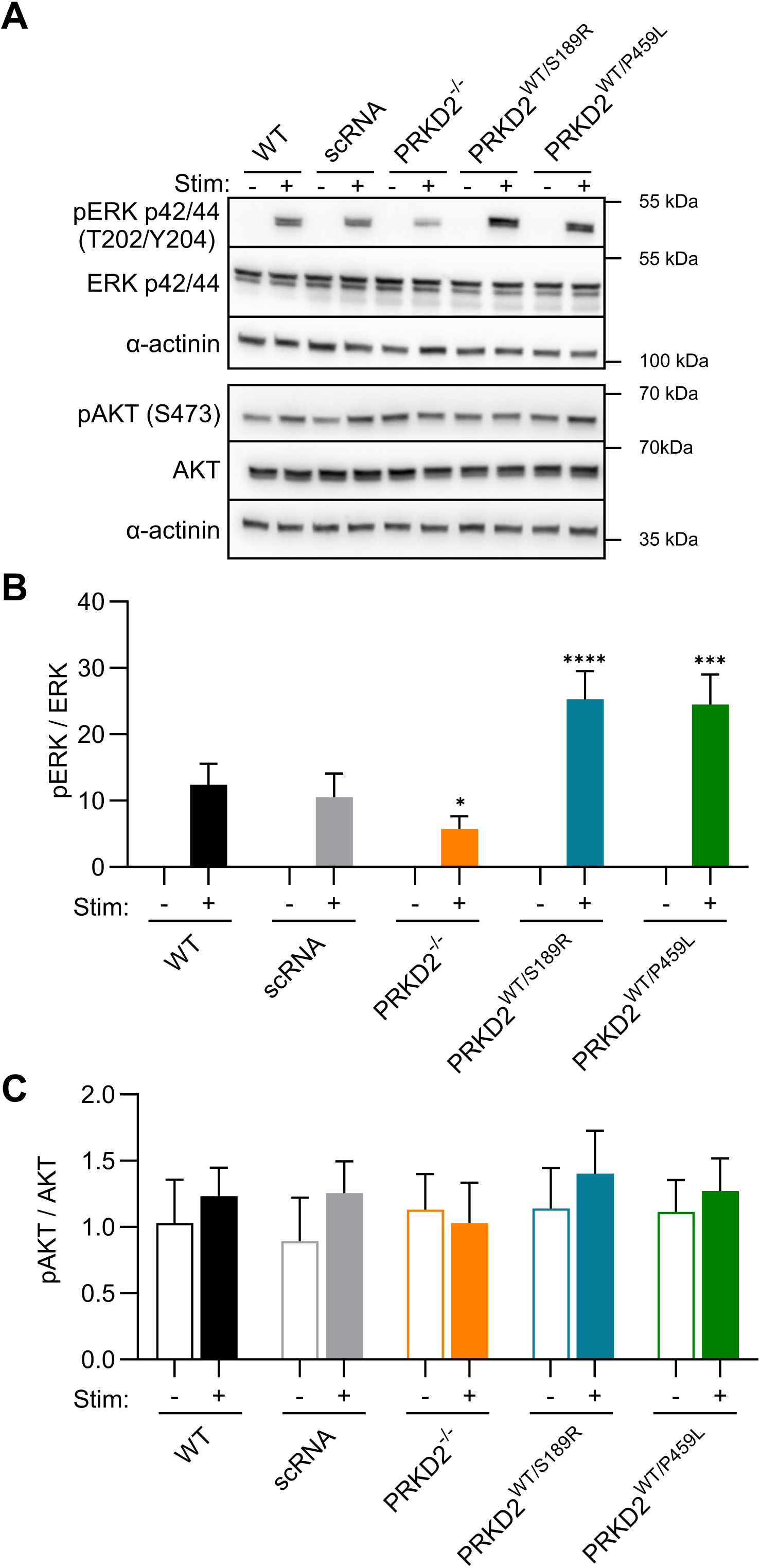
PRKD2 p.Ser189Arg and p.Pro459Leu increase ERK but not AKT phosphorylation in TCR-stimulated KI Jurkats. Jurkat CD4 T cell clones were serum- starved for 24hours in X-VIVO-10 media, then stimulated (+) or not (-) for 30 minutes with anti-CD3 (1µg/mL), anti-CD28 (1µg/mL) and the crosslinker STAR117 (5µg/mL). (**A-C**) Cells were lysed and Western blot performed for the indicated proteins. (**A**) Representative images and (**B-C**) Signal quantitation. (**B**) pERK/total ERK (p42/44), each normalized to α-actinin performed on the same blot. (**C**) pAKT/total AKT, each normalized to GAPDH performed on the same blot. n=3 experiments, each performed on 3 clones per genotype. \**p<0.05*, \*\*\**p<0.001*, \*\*\*\**p<0.0001 vs.* corresponding WT, nested one-way ANOVA with Dunnett’s post-hoc multiple comparisons test.

In summary, both p.Ser189Arg and p.Pro459Leu enhanced TCR-induced NF-κB and ERK activation in heterozygous KI Jurkats, while p.Pro459Leu (Family D) but not p.Ser189Arg (Family A) also increased NFAT activation.

### p.Ser189Arg and p.Pro459Leu increase TCR-mediated activation of heterozygous KI Jurkats

To determine whether increased activation of PRKD2 and downstream signaling pathways translates into altered T cell activation, we measured CD69, PD-1, and CD25 by flow cytometry at 4, 24, and 72 hours of TCR stimulation using αCD3-αCD28 crosslinking (**Fig.6A-B, Supplementary Figure S5**). Both PRKD2^WT/S189R^ and PRKD2^WT/P459L^ cells display significantly increased expression of CD69 and PD-1 at 4 and 24 hours, and CD25 (which appears later) at 24 hours of TCR-stimulation (**Fig.6A-B**). These differences are attenuated by 72 hours of TCR-stimulation (**Supplementary Figure S5**). TCR-stimulated PRKD2-deficient cells show comparable expression of CD69 and PD-1, but reduced expression of CD25 as compared to control cells.

**Figure 6.**
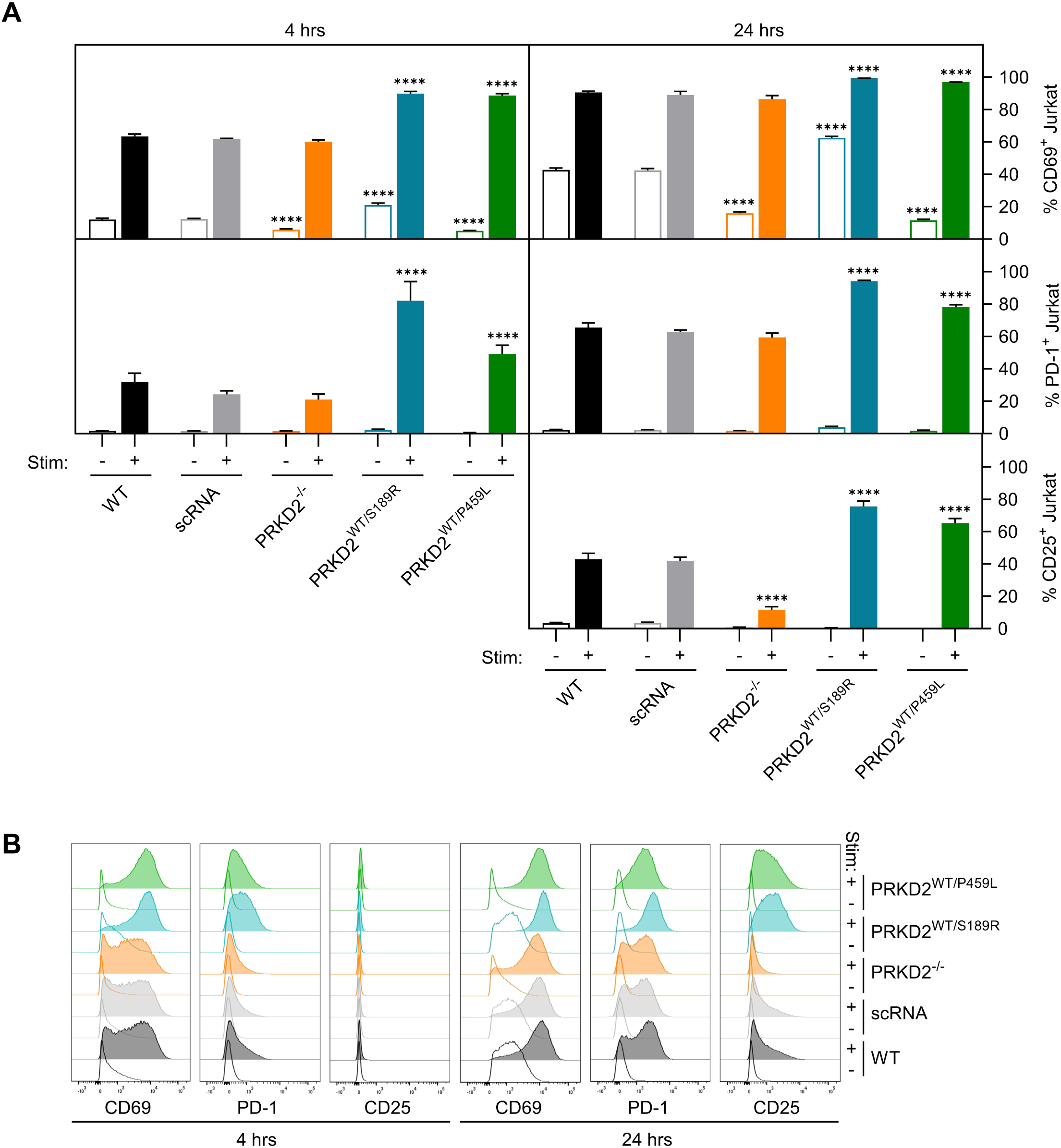
PRKD2 p.Ser189Arg and p.Pro459Leu increase cell surface expression of T cell activation markers in TCR-stimulated KI Jurkats. Jurkat CD4 T cell clones were serum- starved for 24hours in X-Vivo-10 media, then stimulated (+) or not (-) for 4 and 24 hours with anti-CD3 (1µg/mL), anti-CD28 (1µg/mL) and the crosslinker STAR117 (5µg/mL). (**A-B**) Flow cytometry was performed for the T cell activation markers CD69, PD1 and CD25. (**A**) Percentages (CD25 not detected at 4 hours) and (**B**) Representative histograms from n=3 experiments, each performed on 3 clones per genotype. \*\*\*\**p<0.0001 vs.* corresponding WT, nested one-way ANOVA with Dunnett’s post-hoc multiple comparisons test.

For a wider snapshot of the impact of the familial SSc variants, we performed RNASeq on the heterozygous KI, KO, and scRNA Jurkat lines, with or without (4 hours of) TCR stimulation. Principal components analysis (PCA) showed that unstimulated and stimulated PRKD2- deficient cells cluster with their control counterparts, indicating PRKD2 is not required for T cell activation under these conditions (**Fig.7A**). In contrast, PRKD2^WT/S189R^ and PRKD2^WT/P459L^ cells cluster together, away from controls along PC1 (the dominant axis), and apart from each other along PC2 (that accounts for minor additional variability). Gene Set Enrichment Analysis (GSEA) confirmed that multiple T cell activation-related gene-sets are significantly upregulated in PRKD2 KI cells as compared to controls, but remain largely unchanged in KO cells (**Fig.7B**). Along with global T cell activation (**Fig.7C, Supplementary Table S6**), several signaling pathways and transcriptional programs are differentially regulated between KI and control cells, including the NF-κB, JAK/STAT and KRAS pathways (**Fig.7B**). While a subset of these differentially regulated gene-sets is shared between variants, each variant also displays a distinct transcriptional signature (**Fig.7B-C**). These include pathways related to EMT and ECM organization, enriched in PRKD2^WT/S189R^ but not in PRKD2^WT/P459L^ cells. This is intriguing given the extensive cutaneous fibrosis observed in Family A (with the p.Ser189Arg variant) but not in Family D (with the p.Pro459Leu variant).

**Figure 7.**
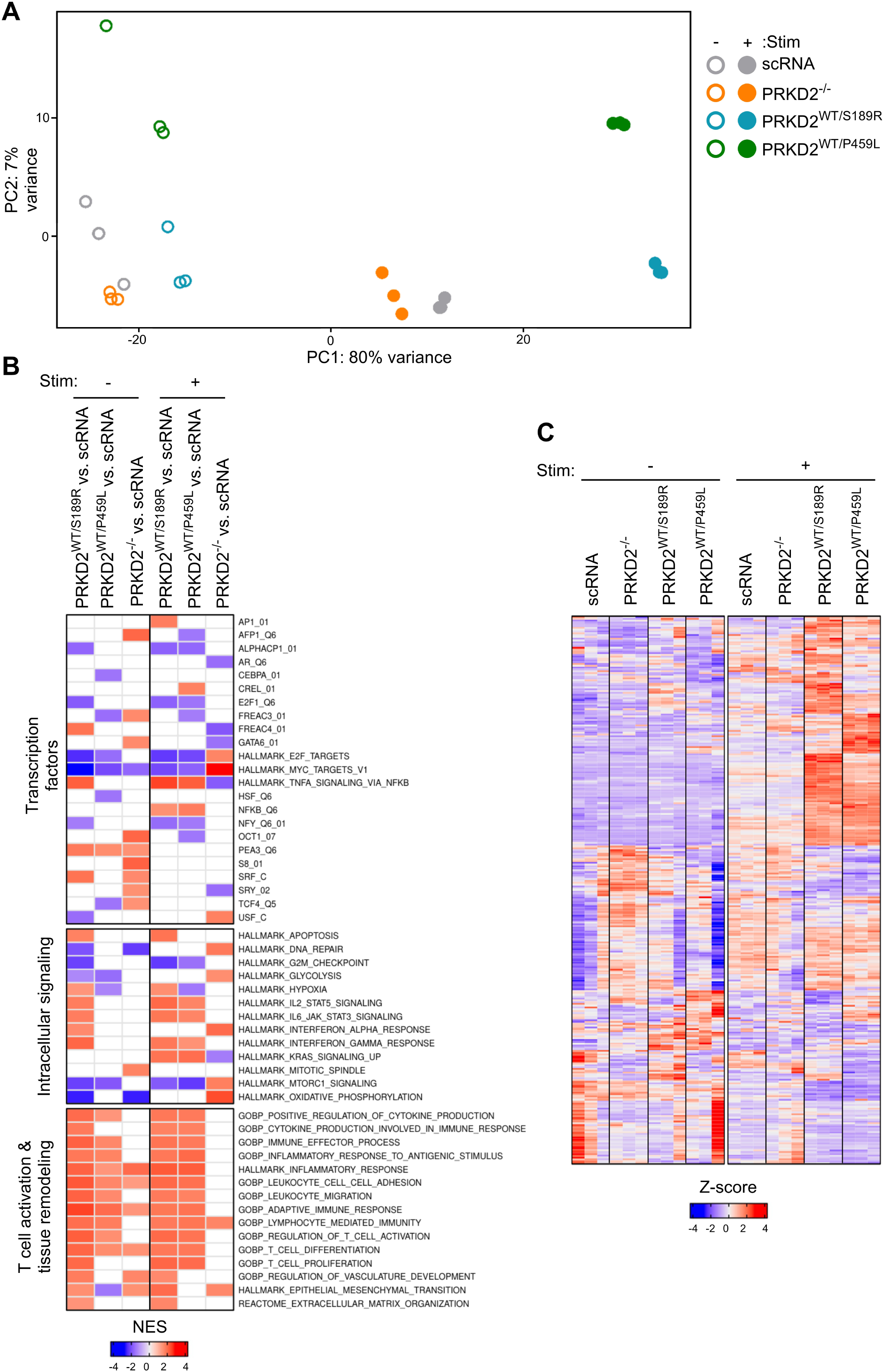
RNASeq and GSEA show higher T cell activation signatures in p.Ser189Arg and p.Pro459Leu KI as compared to scRNA Jurkats. Jurkat CD4 T cell clones were serum- starved for 24hours in X-Vivo-10 media, stimulated (+) or not (-) for 4 hours with anti-CD3 (1µg/mL), anti-CD28 (1µg/mL) and the crosslinker STAR117 (5µg/mL), followed by RNA extraction and and RNASeq. (**A**) Principal Component Analysis (PCA) of RNAseq data from three clones per genotype. Each dot represents one clone: scRNA (grey), PRKD2^-/-^ (orange), PRKD2^WT/S189R^ (blue), PRKD2^WT/P459L^ (green). Unfilled: unstimulated. Filled: TCR-stimulated. (**B**) Heatmap showing normalized enrichment scores (NES) for a selection of significantly enriched gene sets identified by GSEA in the indicated comparisons. (**C**) Heatmap showing the gene-wise scaled values (z-scores) for 313 detected genes from the Regulation_of_T_cell_activation Gene Ontology Biological Process (GOBP) gene-set. Rows: genes. Columns: three clones per genotype per condition.

### Comparable CD4 T cell activation markers in Family A and D patient PBMC *versus* controls, are decreased by a pan-PRKD inhibitor

We next sought to determine whether the increased activation observed in KI Jurkats could be recapitulated in patient-derived cells, and whether T cell activation could be decreased by treatment with a pan-PRKD small-molecule kinase inhibitor.

PBMCs from affected individuals from families A and D, together with two age- and sex-matched controls each, were TCR-stimulated with or without different concentrations of the pan-PRKD inhibitor CRT0066101 (CRT). First, we observed inter-sample variability and no differences in the levels of CD4 T cell activation markers between patients and controls (**Fig.8A-B**). Second, pan-PRKD inhibition strongly decreased T cell activation in all samples at the higher concentrations of CRT tested (5 and 10 µM); treatment with 1 µM had a milder effect (**Fig.8C**). While no differences in CRT “sensitivity” (fold-change versus TCR-stimulated cells) were observed between Family D patient and control samples, a small but consistent effect was observed for Family A: Both A-1 and A-2 showed a somewhat smaller fold-change as compared to their matched controls, across CRT doses and markers (**Fig.8C**).

**Figure 8.**
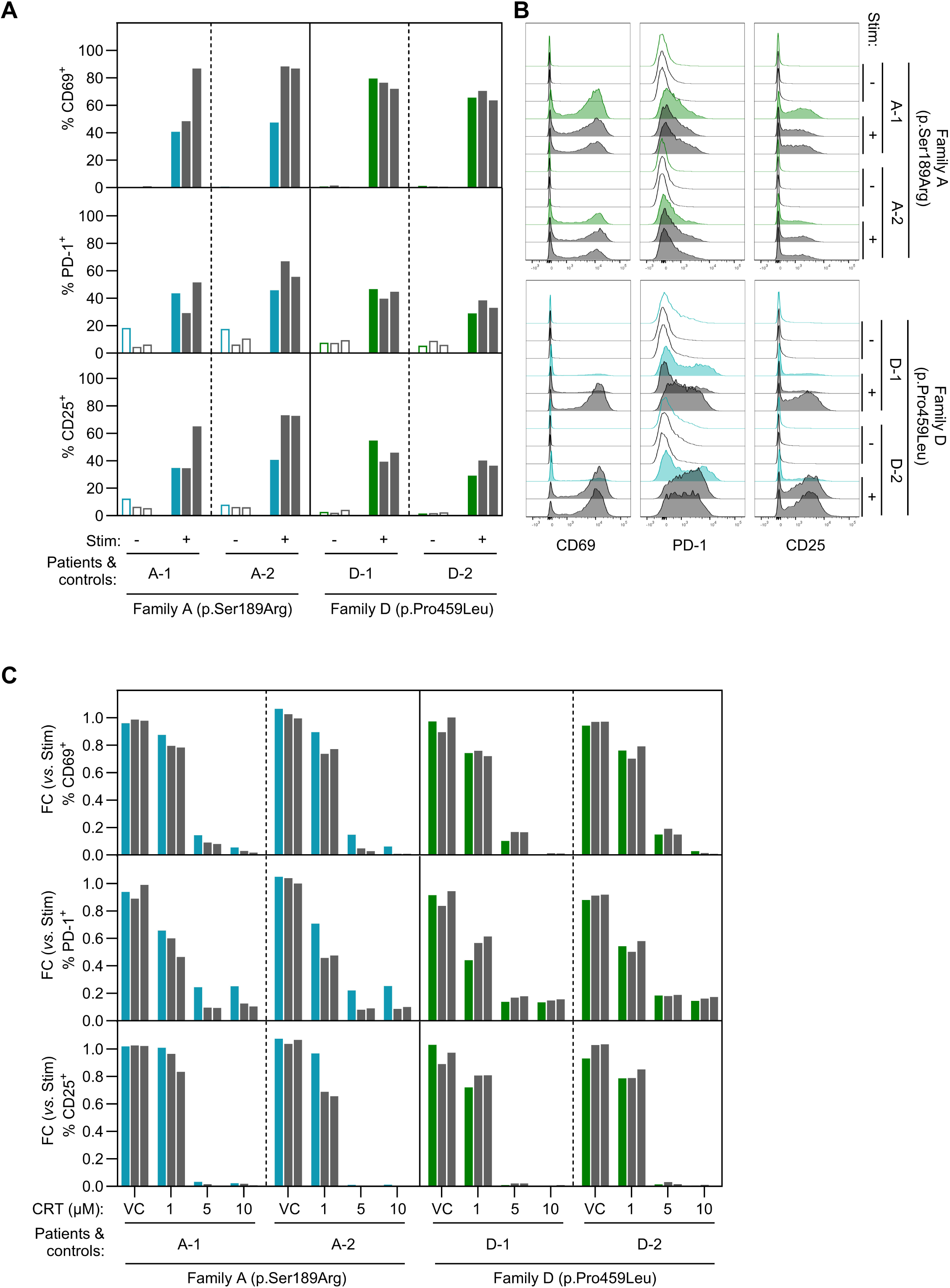
CD4 T cell activation in PBMC from patients in Families A and D *versus* matched controls, and effect of a pan-PRKD inhibitor. PBMC from the affected individuals from Families A (A-1 and A-2, blue) and D (D-1 and D-2, green), and two age- and sex-matched controls each (black) were stimulated with anti-CD3 (1 µg/mL), anti-CD28 (1 µg/mL), STAR117 (5 µg/mL) and IL-2 (50 IU/mL) for 24 hours. Flow cytometry was performed for the T cell activation markers CD69, PD1 and CD25 in CD4-positive cells. **(A)** Bar graphs showing percentages, and **(B)** histograms from unstimulated (clear) and TCR-stimulated (filled) conditions. **(C)** In parallel wells of the same experiment, PBMC were pre-incubated with DMSO as a vehicle control (VC), or different concentrations of a pan-PRKD inhibitor (CRT). T cell stimulation was performed as above, in the presence of (the same concentrations of) DMSO or CRT, for 24 hours. Bar graphs showing fold-changes (FC, stimulation + VC or CRT, versus stimulation alone).

### *PRKD2* rare variant enrichment in sporadic SSc

Analysis of GWAS data from two large cohorts: one French (1,523 cases and 4,581 controls)^35^ and one Japanese (1,428 cases and 112,599 controls)^36^, did not show any significant association of common SNPs in the *PRKD2* locus with SSc (data not shown). In order to assess for (individual and aggregated) enrichment of *PRKD2* coding rare variants in SSc, we performed WES on a series of sporadic SSc patients (n=279, recruited from the three Belgian centers of SSc expertise in the European Reference Network (ERN) ReCONNET: Ciniques universitaires St Luc, Ghent University Hospital, and University Hospitals Leuven) (**Supplementary Tables S7A-C**). Ten additional rare (highest population *MAF* in gnomAD ≤ 0.01) non-synonymous variants were identified, in 17 unrelated individuals. Variant-level association analysis revealed a highly heterogeneous distribution of effect sizes, with odds ratios ranging from near-null or modest effects for more frequent variants (*e.g.* p.Val324Met, p.Glu873Lys), to very strong enrichment signals for ultra-rare variants (*e.g.* p.Arg420Cys, p.Gly493Ala, p.Arg817Gln, p.Arg836His) (**Table 1**). After multiple testing correction, a subset of variants remained significant, with the strongest evidence observed for p.Arg817Gln and p.Arg836His. A Mantel– Haenszel meta-analysis was performed to estimate the overall enrichment of *PRKD2* variants in the cohort relative to gnomAD. Under a fixed-effect model, a statistically significant but modest enrichment was observed in the cohort (OR = 2.44, 95% CI 1.52–3.91; p = 0.0002). However, substantial heterogeneity was observed across variants (Q = 81.89, p < 0.0001; I² = 89.0%), indicating that effect sizes were not uniform. Consistent with this heterogeneity, a random-effects model yielded a substantially larger pooled effect estimate (OR = 22.24, 95% CI 3.47–142.55; p = 0.0011). This suggests that the overall gene-level association is driven by a subset of strongly enriched rare variants rather than a uniform burden across all *PRKD2* sites. WES data from a similarly-sized series (n=285, Ishikawa *et al.*, unpublished) of Japanese patients with sporadic SSc, on the other hand, showed no rare (*MAF* in the gnomAD EAS population ≤ 0.01) coding variants in the *PRKD2* gene.

**Table 1.**
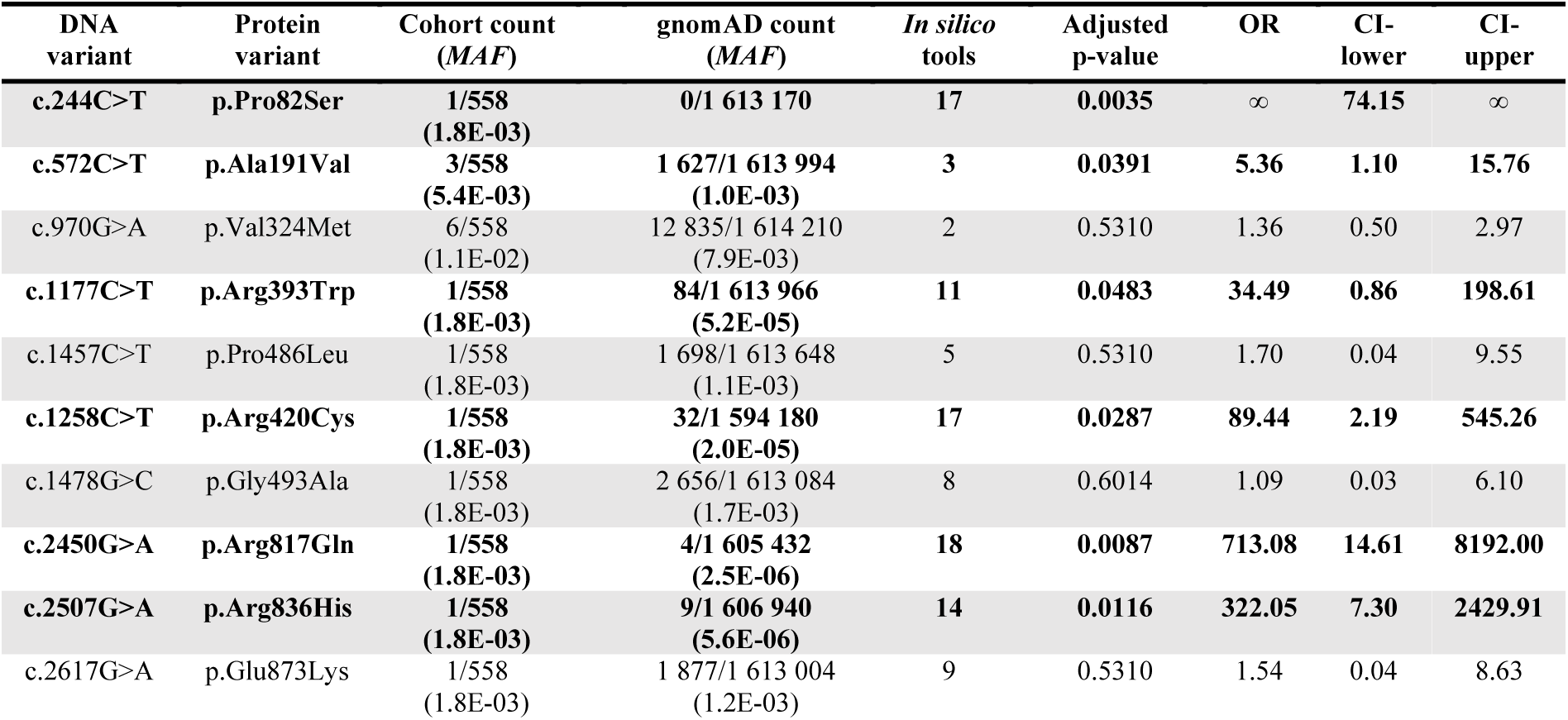
***PRKD2* rare variants in a Belgian cohort of (n=279) sporadic SSc patients.** In bold: variants that show significant enrichment in the SSc cohort *vs* gnomAD. Cohort/gnomAD count: Number of variant alleles/total number of genotyped alleles (*MAF*: Minor Allele Frequency), in the SSc cohort or in gnomAD v4.1.1. *In silico* tools: Number of programs (out of 20) that predict missense variants alter protein function. OR: Odds ratio. CI: Confidence Interval (lower, upper limits).

| DNA variant | Protein variant | Cohort count (MAF) | gnomAD count (MAF) | *In silico* tools | Adjusted p-value | OR | CI-lower | CI-upper |
| --- | --- | --- | --- | --- | --- | --- | --- | --- |
| c.244C>T | p.Pro82Ser | 1/558<br>(1.8E-03) | 0/1 613 170 | 17 | 0.0035 | ∞ | 74.15 | ∞ |
| c.572C>T | p.Ala191Val | 3/558<br>(5.4E-03) | 1 627/1 613 994<br>(1.0E-03) | 3 | 0.0391 | 5.36 | 1.10 | 15.76 |
| c.970G>A | p.Val324Met | 6/558<br>(1.1E-02) | 12 835/1 614 210<br>(7.9E-03) | 2 | 0.5310 | 1.36 | 0.50 | 2.97 |
| c.1177C>T | p.Arg393Trp | 1/558<br>(1.8E-03) | 84/1 613 966<br>(5.2E-05) | 11 | 0.0483 | 34.49 | 0.86 | 198.61 |
| c.1457C>T | p.Pro486Leu | 1/558<br>(1.8E-03) | 1 698/1 613 648<br>(1.1E-03) | 5 | 0.5310 | 1.70 | 0.04 | 9.55 |
| c.1258C>T | p.Arg420Cys | 1/558<br>(1.8E-03) | 32/1 594 180<br>(2.0E-05) | 17 | 0.0287 | 89.44 | 2.19 | 545.26 |
| c.1478G>C | p.Gly493Ala | 1/558<br>(1.8E-03) | 2 656/1 613 084<br>(1.7E-03) | 8 | 0.6014 | 1.09 | 0.03 | 6.10 |
| c.2450G>A | p.Arg817Gln | 1/558<br>(1.8E-03) | 4/1 605 432<br>(2.5E-06) | 18 | 0.0087 | 713.08 | 14.61 | 8192.00 |
| c.2507G>A | p.Arg836His | 1/558<br>(1.8E-03) | 9/1 606 940<br>(5.6E-06) | 14 | 0.0116 | 322.05 | 7.30 | 2429.91 |
| c.2617G>A | p.Glu873Lys | 1/558<br>(1.8E-03) | 1 877/1 613 004<br>(1.2E-03) | 9 | 0.5310 | 1.54 | 0.04 | 8.63 |

## Discussion

We applied a rare disease genetics approach to familial SSc, in order to identify and functionally test candidate gene variants with potential disease-predisposing effects. WES identified >250 candidate genes (with disease co-segregating rare variants, predicted to affect protein function by multiple *in silico* tools) across the six families, none shared by more than two. This degree of genetic heterogeneity is consistent with the phenotypic heterogeneity observed among the families, and indeed in SSc more broadly. Similar patterns are observed in systemic lupus erythematosus (SLE): a clinically heterogeneous, multi-system autoimmune disease that is typically complex and rarely familial. More than 30 genes have been identified to cause monogenic (lupus, lupus-like) forms^37–39^. Together, they account for a small fraction of cases, and encompass a diversity of phenotypes. They nevertheless shed light on key pathogenic mechanisms and pathways in SLE. The genetic analysis of familial SSc, in contrast, remains under-utilized. With 51 families described thus far across six studies^40–45^, genetic analysis has to our knowledge been reported for only one (WES; with no functional testing of candidate gene variants^41^). Besides this, in a 2021 study, Lepelley *et al.* reported SSc in two unrelated patients (out of seven) with a cGAS/STING-activating mutation in the *ATAD3A* gene, which causes rare, severe neurological phenotypes^46^. We did not identify (novel or previously reported) *ATAD3A* rare variants in the current series.

Notwithstanding the putative genetic heterogeneity of familial SSc, we made the deliberate choice, in this first instance, of prioritizing a “shared” gene for functional testing. Aside from meeting this objective criterion, *PRKD2* is an attractive candidate: knock-out mice display polyclonal hypergammaglobulinemia, with an increase in T follicular helper (T_FH_) CD4 T cells, and a concomitant increase in germinal centers (GCs), GC B cells, plasma cells, T-dependent B cell responses and autoantibody production^47^. Replacement of PRKD2 with a catalytically defective (mouse Ser707Ala-Ser711Ala (PRKD2^SSAA^)) version, on the other hand, seems to *decrease* antigen-specific T-dependent B cell responses in knock-in mice^15, 48^, suggesting kinase-dependent and -independent roles for the protein. PRKD2^SSAA^ knock-in mice are also protected against bleomycin-induced skin fibrosis, with PRKD2 expression restricted mainly to infiltrating macrophages in the affected area^49^. Given the established role of PRKD2 in T cell function, and the autoimmune phenotype observed in both families (one with and one without significant skin fibrosis), we studied the effects of the variants in an isogenic human CD4 T cell model.

Chronically hyperactivated T cells and disease-specific autoantibodies are central features of SSc, with affected tissues showing prominent immune cell infiltration, particularly by CD4 T cells^50–52^. T_H2_, T_H17_ and T_FH_ cells are found to be elevated in SSc patients as compared to healthy controls, whereas Tregs, while increased in number, seem to exhibit impaired immunosuppressive function^53–60^. We demonstrate overlapping yet distinct activating effects of both *PRKD2* variants in Jurkat T cells. Key differences include increased TCR-induced activation of NFAT (by p.Pro459Leu but not p.Ser189Arg), and the nuclear translocation of PRKD2 (increased by p.Ser189Arg, decreased by p.Pro459Leu). It is challenging to decipher the contributions of NF-κB (modulated by both variants), NFAT, and the combination thereof, to different aspects of T cell differentiation and function. Studies using pathway inhibitors suggest that certain transcriptional targets are responsive to one or the other, while many require both^61^. Data from inborn errors of immunity (IEI) suggest that overactivation of NF-κB (*e.g.* due to *TNFAIP3* or *TNIP1* loss, *RELA* gain of function) can cause autoinflammatory/autoimmune phenotypes, in some cases with T cell exhaustion or anergy^62^. Overactivation of both NFAT and NF-κB in T cells, for example due to variants in TCR-proximal adaptors such as *PLCG2* or *ZAP70*, also results in immune dysregulation and autoinflammation^63, 64^. To our knowledge, no *PRKD2* variants have been described in IEI, nor indeed in other inherited human disorders.

Nuclear-cytoplasmic distribution of active PRKD2 protein can also impact T cell differentiation and function. Studies in mice and human cells have shown that the cytoplasmic fraction of active PRKD2 has an important role in binding and phosphorylating the master T_FH_ transcription factor BCL6, limiting its nuclear translocation^47^. In the absence of PRKD2 protein, nuclear entry of BCL6 is increased, resulting in excessive T_FH_ differentiation and T-dependent B cell responses. The nuclear fraction of active PRKD2, on the other hand, phosphorylates HDAC7, promoting its nuclear export and subsequent expression of genes including *IRF4* (that promotes T_H2_ differentiation) and *NR4A1* (Nur77, important in thymic selection)^65–72^. Retention of HDAC7 in the nucleus increases thymocyte escape from negative selection, resulting in increased autoreactive T cells in the periphery^65–67^. Based on these observations (**Supplementary Figure S6**), it is possible that p.Ser189Arg, which increases nuclear translocation of PRKD2, permits increased nuclear entry of BCL6, promoting T_FH_ differentiation. p.Pro459Leu, which decreases nuclear translocation, may instead favor T_H2_ differentiation. Thus, each may recapitulate certain aspects of PRKD2 protein loss, despite globally increasing its activation. While such hypothesized (but unproven) T cell effects may contribute to the phenotypic differences between the two families, differential effects of these variants in other cell types (*e.g.* macrophages) may also play an important role. These may act in concert with stochastic and/or somatic (epi)genetic effects, polygenic risk score (PRS), and/or the effects of other strong candidate genes in a di/oligogenic model of disease inheritance.

We report ten additional heterozygous, missense rare variants in *PRKD2*, six of which show significant enrichment in a Belgian sporadic SSc series as compared to the gnomAD population. Together, they suggest significant gene-level rare variant “burden” for *PRKD2* in SSc (not captured by traditional GWAS), with highly heterogeneous effect sizes that bear substantiation in larger (WES, WGS) SSc cohorts. Intriguingly, WES data from a Japanese sporadic SSc series would suggest that any such association of *PRKD2* rare variants may differ between populations, as is also often the case with susceptibility alleles identified by GWAS.

In the first study of its kind in SSc, we provide genetic and functional *in vitro* evidence for *PRKD2* as a candidate susceptibility gene. Complementary studies in additional human cell-types and in animal models, will be valuable in validating its pathogenic potential. The full list of candidate gene variants (meeting filtering criteria, from all six families) provided here, also opens the door to the testing of additional candidates, including but not limited to potential “partner” genes that modulate the effects of *PRKD2* in affected families.

## Materials & methods

### Patient samples

These studies were approved by the ethics committees of Cliniques universitaires St Luc (2015/17NOV/629, 2025/10FEV/060), Ghent University Hospital (EC/2016/0209) and University Hospitals Leuven (S52057-ML5780). Patients fulfilled ACR/EULAR classification criteria for SSc, and stratified into lSSc, lcSSc and dcSSc subsets according to LeRoy and Medsger^73–75^. SSc-related organ involvement was evaluated as previously described in^76^. French (1,523 cases and 4,581 controls, including previously published data^35^ and the Three-City Study cohort for controls) and Japanese (1,428 cases and 112,599 controls^36^) GWAS datasets were used to assess for common-variant association of the *PRKD2* locus with sporadic SSc.

### Whole exome sequencing and variant filtering

Genomic DNA was extracted from whole blood using the Wizard Genomic DNA extraction kit (Promega, #A1620). Whole exome sequencing (WES) was performed at Macrogen, Europe (Illumina HiSeq4000 with the SureSelect v6 capture kit: familial SSc samples; NovaSeq X with the Twist Exome 2.0 capture kit: sporadic SSc samples). Raw data was processed using a standard pipeline (**Supplementary Methods**). Candidate variants shared by both affected members within families were retained if they met the following criteria: (a) PASS standard GATK quality control filters, (b) Highest population minor allele frequency (popmax *MAF*) ≤0.001 in gnomAD v4.1.1 and in the local WES database (PGEN Genomics platform, UCLouvain), (c) Consensus splice-site changes, frame-shifting variants, nonsense substitutions, or missense variants predicted to have a deleterious effect by ≥5 out of 20 *in silico* tools (see **Supplementary Methods**).

### Jurkat cell culture

Jurkat E6.1 cells were cultured at 37°C with 5% CO_2_, in RPMI medium supplemented with Glutamax (Gibco, #72400047), 0.1% 2-mercaptoethanol (Gibco, #21985023), 1% Non-Essential Amino Acids Solution (Gibco, #11140035), 1% Antibiotic-Antimycotic (Gibco, #15240062), and 10% decomplemented Foetal Bovine Serum (FBS) (VWR, #HYCLSV30160.03).

### Generation of knock-out (KO) and knock-in (KI) Jurkat clones

CRISPR RNAs (crRNAs, designed using the Integrated DNA Technologies (IDT) online tool) and trans-activating crRNA (tracrRNA, IDT, #1072532) were purchased from Integrated DNA Technologies (IDT). To generate KO Jurkat cells, crRNA and tracrRNA (100 µM each) were mixed and annealed by heating at 95°C for 5 minutes, followed by gradual cooling from 95°C to 25°C in 5°C decrements every 2 minutes. 3µL of the crRNA-tracrRNA duplex was incubated with 2µL of recombinant Cas9 protein (ThermoFisher, #A36499) for 15 minutes at room temperature. 1x10^6^ Jurkat cells were resuspended in 20µL SE Cell Line 4D Nucleofection Solution, gently mixed with the crRNA-tracrRNA-Cas9 complex, and nucleofected in a Nucleocuvette Vessel (Lonza, #V4XC-1032) using the CL-120 program on a 4D-Nucleofector X Unit. Cells were allowed to recover in culture medium for a minimum of 48 hours, followed by single-cell sorting on a MA900 Cell Sorter (Sony, #MA900). Clones were screened by Western blot for loss of PRKD2 protein, and PCR-sequencing for the presence of insertions-deletions in exon 1 of the *PRKD2* gene.

For the generation of KI cells, a similar nucleofection protocol was used, with the addition of 1.2µL of Electroporation Enhancer (IDT, #1075915; 100µM) and 1.2µL HDR Donor Oligo (100µM, designed and ordered from IDT). After nucleofection, cells were incubated for 16 hours in culture medium containing 1.7µL/mL HDR Enhancer V2 (IDT, #10007910), then washed, allowed to recover for two weeks in culture medium, and single-cell sorted by FACS. All crRNA and HDR Donor Oligo sequences are listed in **Supplementary Methods**.

Clones were expanded (2x96 well plates per variant) and screened for presence and heterozygosity of variants by targeted Next Generation Sequencing (NGS) of exons 5 and 11 (**Supplementary Methods**). Three clones per genotype were further expanded for use across all experiments.

### Lentiviral transduction of NFAT and NF-κB reporter constructs

Lentiviral particles were produced by transient transfection of HEK293T cells with a reporter construct encoding GFP under the control of an NFAT- or NF-κB-responsive element and mCherry under a constitutive PGK promoter (kind gift from Prof. Pierre Coulie, de Duve Institute, UCLouvain), together with the packaging plasmids pGag-Pol, pVSV-G, and pRev, using the calcium phosphate transfection method. Viral supernatants were collected 48 hours post-transfection, filtered through a 0.45µM filter, concentrated with Vivaspin20 (Sartorius, #VS2022) and stored at -80°C. Jurkat cells were seeded at 2x10^5^ cells/mL and incubated with viral supernatants and 10µg/mL polybrene (Millipore, #TR-1003-G) for 72 hours. Cells were sorted by FACS (Sony, #MA900) to select populations with equivalent mCherry MFI across cell lines.

### TCR stimulation of Jurkat clones

In order to maintain uniformly low basal T cell activation, all clonal cell-lines were cultured at comparable cell densities (<5x10^5^ cells/mL). Cells were serum-starved for 24 hours in X-Vivo-10 media (Lonza, #BE04-380Q) with 1% Antibiotic-Antimycotic, then stimulated using 1µg/mL anti-human CD3 antibody (BioLegend, #317326), 1µg/mL anti-human CD28 antibody (BioLegend, #302934) and 5µg/mL goat anti-mouse IgG (BioRad, #STAR117) for the times indicated per experiment.

### TCR stimulation of primary peripheral blood mononuclear cells (PBMC)

PBMC were isolated from blood collected in EDTA tubes by Ficoll-Paque density gradient centrifugation (VWR, #17-5446-53) according to manufacturer’s protocol, and cryopreserved at -80°C in FBS containing 10% DMSO (Sigma-Aldrich, #D2650) until use. PBMCs were stimulated at 5x10^5^ cells/mL in RPMI medium supplemented with Glutamax (Gibco, #72400047), 0.1% 2-mercaptoethanol (Gibco, #21985023), 1% Non-Essential Amino Acids

Solution (Gibco, #11140035), 1% Antibiotic-Antimycotic (Gibco, #15240062) and 10% decomplemented Foetal Bovine Serum (FBS) (VWR, #HYCLSV30160.03), supplemented with 50IU/mL recombinant IL-2 (ThermoFisher, #HZ-1015). Cells were pre-incubated for 1 hour in the same medium containing the pan-PRKD inhibitor CRT0066101 (Sanbio, #HY-15698-5) at the indicated concentrations, then stimulated with 1µg/mL anti-human CD3 antibody (BioLegend, #317326), 1µg/mL anti-human CD28 antibody (BioLegend, #302934) and 5µg/mL goat anti-mouse IgG (BioRad, #STAR117).

### Cell lysis and Western blot

1x10^6^ cells were lysed in 100µL of lysis buffer (Tris-HCl 50mM, NaCl 150mM, EDTA 2mM, NP40 0.1%, Complete (Roche, #11873580001, one tab/20mL), PhosSTOP (Roche, #4906845001, one tab/10mL)). Lysates were incubated for 20 minutes at 4°C on a shaker, sonicated for 15sec at 10% amplitude, and centrifuged at 12000g for 10 minutes at 4°C to remove debris. Lysates were mixed with 1x Lane Marker Reducing Sample Buffer (ThermoFisher, #39000), heated for 10 minutes at 95°C, and separated by SDS-PAGE, followed by transfer onto PVDF membranes for Western blot (**Supplementary Methods**).

### Immunofluorescence

5x10^5^ Jurkat cells were centrifuged at 1000g for 1 minute onto glass coverslips (VWR, #630- 1597) coated with Poly-L-Lysine (Sigma, #25988-63-0). Cells were fixed with pre-warmed (37°C) 4% paraformaldehyde (Sigma, #252549) for 30 minutes at room temperature, washed 3x5 minutes with PBS, and permeabilized with PBS-0.5% Triton X-100 for 15 minutes at room temperature. After blocking in PBS-0.5% Triton X-100-5% BSA-2% milk for 1.5 hours at room temperature, cells were incubated overnight at 4°C in anti-PRKD2 primary antibody (ThermoFisher, #PA5-82764) diluted 1:200 in blocking buffer. Coverslips were washed 5x5 minutes with PBS-0.5% Triton X-100 and incubated for 1 hour at room temperature with AF488-conjugated donkey anti-rabbit IgG secondary antibody (Sanbio, #711-545-152) diluted 1:200 in blocking buffer. After 5x5 minute washes with PBS-0.5% Triton X-100 and one wash of 5 minutes with diH_2_O, coverslips were mounted on microscope slides (ThermoFisher, #J100AMNZ) using mounting medium containing DAPI (Abcam, #ab104139). Slides were dried for 30 minutes at room temperature and stored at -20°C until imaging on an LSM980- multiphoton confocal microscope (Zeiss).

### Immunofluorescence data analysis

PRKD2 signal quantitation was performed using an R script. Nuclear segmentation was based on DAPI staining. Cell segmentation was based on PRKD2 (AF488) staining, and the cytoplasm of each cell was defined as the cell-segmented area, excluding its nuclear area. The nuclear/cytoplasmic (N/C) ratio was calculated by dividing the mean nuclear PRKD2 fluorescence intensity by the mean cytoplasmic PRKD2 fluorescence intensity, for each cell. Three independent experiments were performed, each on three clones per genotype per condition, each with 3-5 non-overlapping images collected from the same slide. A total of 1050±47 cells (120±10 cells x 3 clones tested x 3 experiments) per genotype per condition were quantified.

### Flow cytometry

Cells were blocked for 10 minutes at room temperature with Human TruStain FcX (BioLegend, #422302), then incubated for 20 minutes at 4°C with fluorescence-conjugated antibodies (**Supplementary Methods**) diluted in PBS-1mM EDTA-1% FBS. Cells were washed 3x and resuspended in the same buffer, with the viability dye DRAQ7 (1:1000, BioStatus, #DR71000). Data were acquired on a BD LSR Fortessa^TM^ and analyzed using FlowJo v10.10.

## Calcium flux assay

3x10^6^ cells were serum-starved for 24 hours in X Vivo-10 medium at 1x10^6^ cells/mL. Cells were then incubated with Fluo-4 AM (ThermoFischer, #F14217) diluted at 0.2µM in X-Vivo-10 for 20 minutes at 37°C, washed twice and resuspended in X-Vivo-10 containing DRAQ7 (1:1000). Baseline fluorescence was recorded for 60 seconds, after which cells were stimulated with 1µg/mL anti-CD3, 1µg/mL anti-CD28 and 5µg/mL goat anti-mouse IgG as described above. Samples were gently mixed, returned to the cytometer, and acquired for an additional 9 minutes. Data were acquired on a BD LSR FortessaTM and analyzed using FlowJo v10.10.

## RNASeq

RNA was extracted from cells using Tripure isolation reagent (Merck, #11667165001) according supplier’s protocol. RNA quantity and quality were measured using the 4150 TapeStation System (Agilent). TruSeq stranded mRNA library preparation and RNASeq were performed at Macrogen Europe. Raw data were processed using a standard RNAseq pipeline for read aligment to the human genome (GRCh38), evaluation of gene expression, and differential expression and Gene Set Enrichment pathway analysis (**Supplementary Methods**).

## Variant-level and gene-level enrichment analyses of sporadic SSc WES

We evaluated the enrichment of rare coding *PRKD2* variants in the study cohort compared with the gnomAD reference population using allele-count–based association testing. For each variant, a 2×2 contingency table was constructed using alternate and total allele counts in the cohort (n = 279 individuals; 558 alleles) and gnomAD. Variant-level effect sizes were estimated as odds ratios (ORs) with corresponding 95% confidence intervals (CIs) using Fisher’s exact test. To assess the overall gene-level association while accounting for between-variant variability, we performed a Mantel–Haenszel meta-analysis across all variants using the meta R package (metabin). Both fixed-effect (common-effect) and random-effects models were computed. The random-effects model used a restricted maximum likelihood (REML) estimator for between-study variance (τ²). Heterogeneity between variants was quantified using Cochran’s Q statistic, the I² statistic, and τ² estimates. Continuity correction (0.5) was applied for zero-cell counts. Multiple testing correction across variants was performed using the Benjamini–Hochberg procedure.

## Statistical analyses

Except where specified above, all statistical analyses were performed on GraphPad Prism 10. Tests used and p-values are detailed in each Figure legend.

## Data availability statement

Ethics regulations in Belgium do not allow sharing of the germline WES data. The genomic coordinates of the variants identified in this study, according to GRCh38, are provided in **Supplementary Table S4**. RNASeq data from Jurkat cells are available on the GEO (NCBI) repository (accession number GSE346025). All other data are provided in the article and its supplementary materials.

## Data Availability

Ethics regulations in Belgium do not allow sharing of the germline WES data. The genomic coordinates of the variants identified in this study, according to GRCh38, are provided in Supplementary Table S4. RNASeq data from Jurkat cells are available on the GEO (NCBI) repository (accession number GSE346025). All other data are provided in the article and its supplementary materials.

## Acknowledgements

We are grateful to patients and their families for their participation. The authors thank Patrick Vandersmissen for technical assistance with immunofluorescence and confocal microscopy, Nicolas Dauguet for technical assistance with flow cytometry and cell sorting, and Melissa De Decker and Tatiana Sokolova for logistical coordination and database management. We thank Bernard Lauwerys, Frédéric Houssiau and Pierre Coulie for helpful discussions and critical reading of the manuscript. Nisha Limaye is a Senior Research Associate of the Fonds de la Recherche Scientifique (FNRS), Belgium. Vanessa Smith is a founding and steering committee member of the European Reference Network on Rare Connective Tissue and Musculoskeletal Diseases (ERN ReCONNET), and a Senior Clinical Investigator of the Research Foundation- Flanders (FWO) (1802925N).

## Supplementary materials

**Supplementary Table S1.** Clinical description and case histories of the affected members of six SSc families.

**Supplementary Table S2.** Summary of 51 previously published SSc families, summarized _from_40-45_._

**Supplementary Table S3.** Number of variants, per family, retained after each filtering step.

**Supplementary Table S4.** Lists of all variants that passed WES filtering criteria, per family.

**Supplementary Table S5.** Genes with variants that passed filtering criteria in more than one family.

**Supplementary Table S6.** Gene-wise scaled values (z-scores) for 313 detected genes from the Regulation_of_T_cell_activation Gene Ontology Biological Process (GOBP) gene-set.

**Supplementary Tables S7A-C.** Summary of clinical characteristics of the sporadic SSc series.

**A.** All patients, **B.** Subset of patients with rare coding variants in *PRKD2*, **C.** Subset of patients with rare, coding variants found to be statistically enriched in the series as compared to gnomAD v4.1.1.

**Supplementary Methods.**

**Supplementary Figure S1.** Full-length cDNA PCR and Sanger sequencing of *PRKD2* from patient PBMCs.

**Supplementary Figure S2.** Schematic representation of TCR-induced PRKD2 activation and function.

**Supplementary Figure S3.** p.Ser189Arg and p.Pro459Leu do not alter ZAP70 and LAT phosphorylation upon TCR stimulation of heterozygous KI Jurkats.

**Supplementary Figure S4.** p.Ser189Arg and p.Pro459Leu show different kinetics of TCR-stimulated PRKD2 autophosphorylation in heterozygous KI Jurkats.

**Supplementary Figure S5.** Effects of p.Ser189Arg and p.Pro459Leu on expression of T cell activation markers at 72 hours of TCR-stimulation of heterozygous KI Jurkats.

**Supplementary Figure S6.** Schematic representation of the role of active PRKD2 protein in the cytoplasmic and nuclear compartments in T cells.

