## Supplementary Figures S1-S6 for "Activating variants in PRKD2 in two families with Systemic sclerosis, and enrichment of rare variants in a sporadic series"

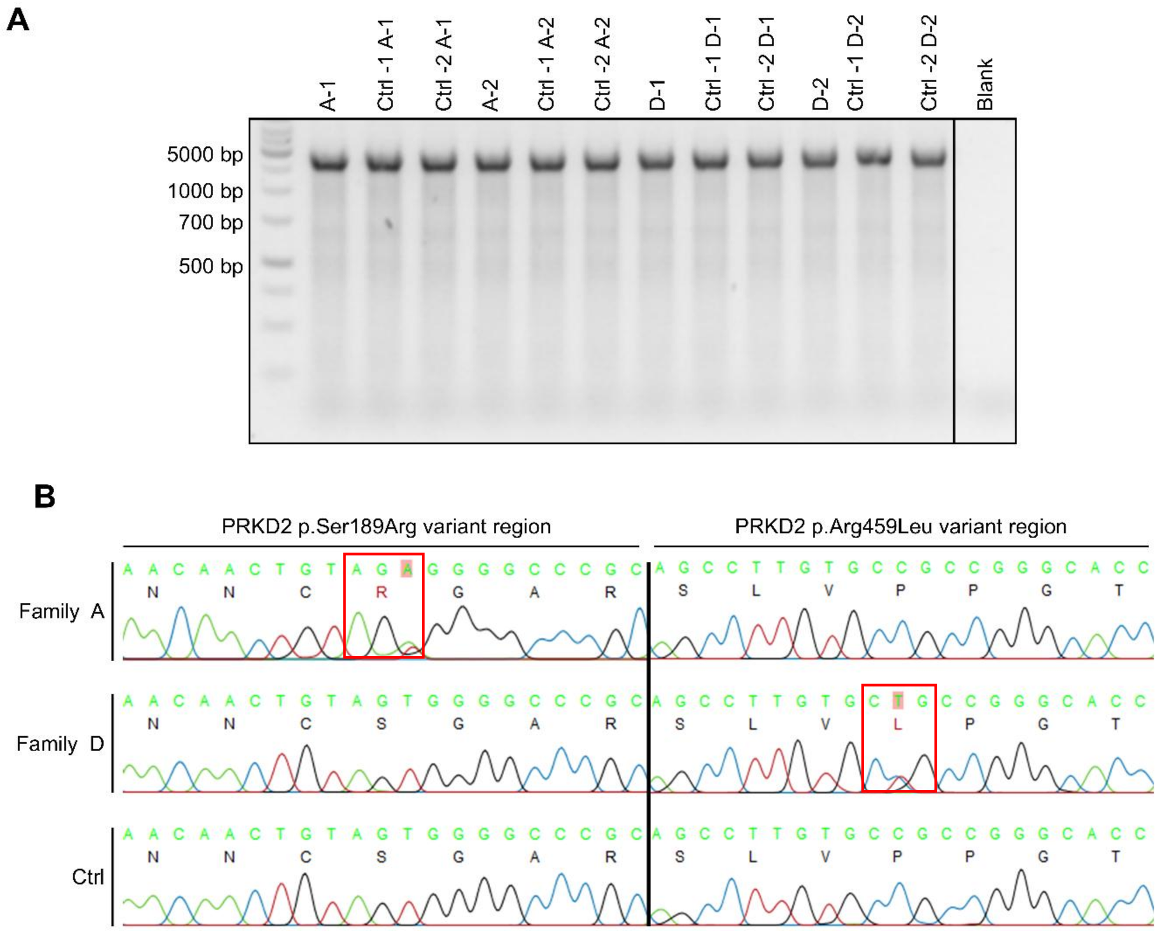


**Supplementary Figure S1: Full-length cDNA PCR and Sanger sequencing of *PRKD2* from patient PBMCs.** (**A**) Agarose gel electrophoresis showing a single band of the expected size, amplified from PBMC cDNA from patients and controls. (**B**) Representative Sanger sequencing chromatograms of column-purified PCR products, showing heterozygous (left: T>A, right: C>T) variant-peaks in patient samples. A-1, A-2 and D-1, D-2: Affected members of Families A and D, respectively. Ctrl-1 and Ctrl-2: Corresponding age- and sex-matched controls.

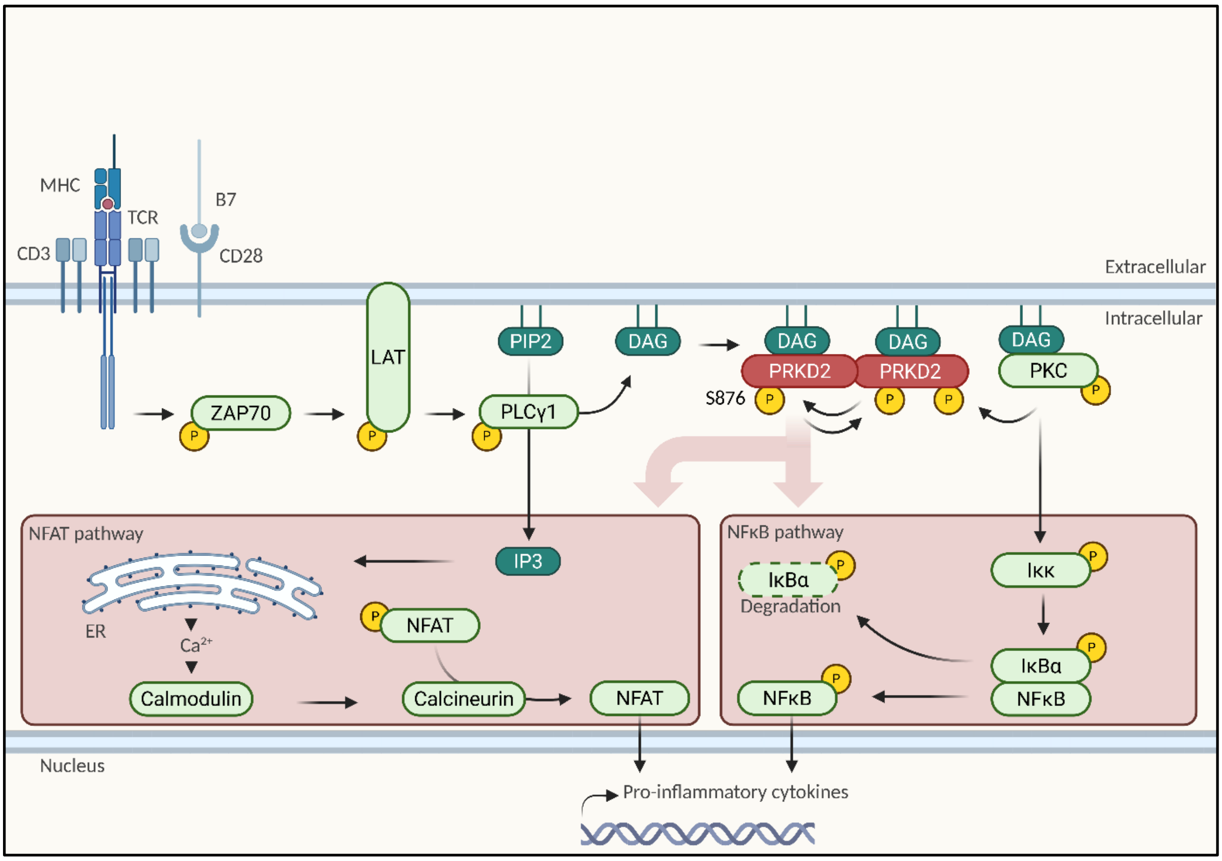


**Supplementary Figure S2: Schematic representation of TCR-induced PRKD2 activation and function.** Upon recognition of a specific MHC-peptide complex by the TCR, or by αCD3/αCD28 antibody crosslinking, ZAP70 is recruited and activated by the ζ chains of the CD3 complex, allowing for activation of LAT. This leads to the activation of PLCγ1, which hydrolyzes PIP2 (Phosphatidylinositol 4,5-bisphosphate) into DAG (Diacylglycerol) and IP3 (Inositol triphosphate). IP3 mobilizes $Ca^{2+}$ from the endoplasmic reticulum, activating calmodulin and calcineurin, which dephosphorylates NFAT, allowing for its nuclear translocation. Simultaneously, DAG activates PKC and recruits PRKD2 to the plasma membrane, allowing for its dimerization and auto trans-phosphorylation on Ser876. Phosphorylation of PRKD2 on Ser706/710 by PKC is further required for full activation. PKC also activates Iκκ, leading to phosphorylation and degradation of IκBα, allowing for NF-κB release and nuclear translocation. PRKD2 regulates both NFAT and NF-κB pathways via unknown mechanisms. The nuclear translocation of the transcription factors NFAT and NF-κB leads to the production of different sets of transcripts including cytokines, critical in determining the degree and type of T cell responses induced.

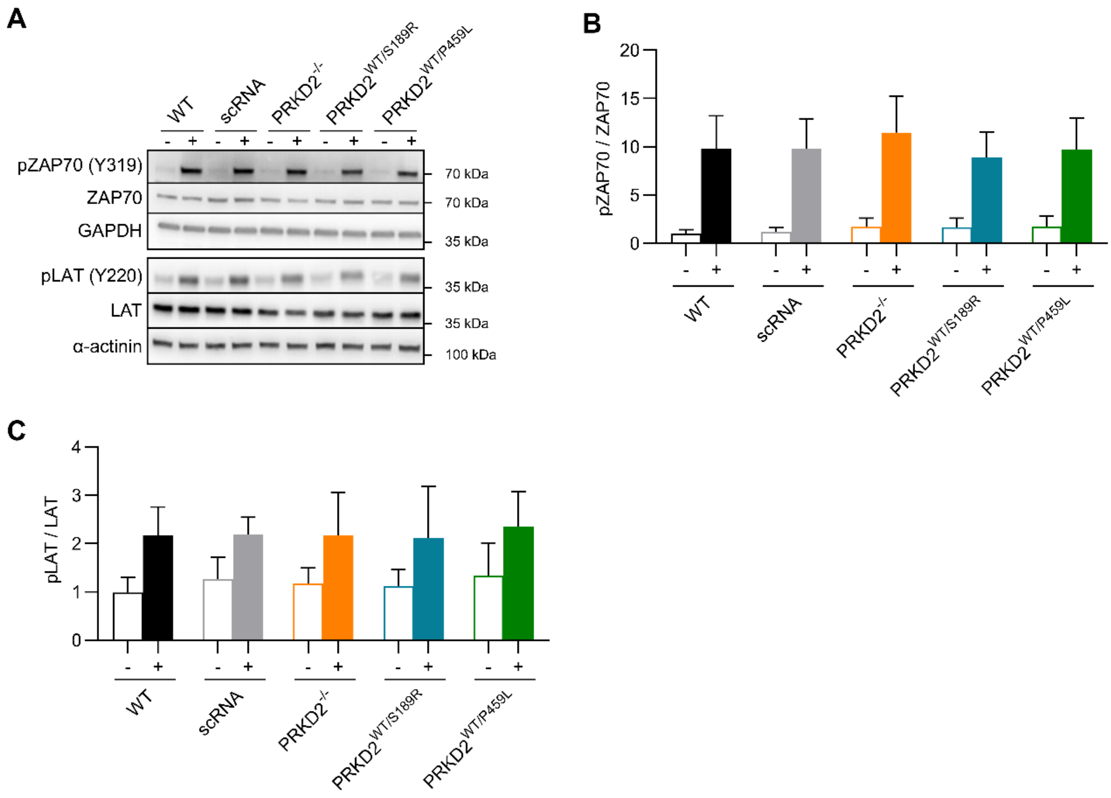


**Supplementary Figure S3: p.Ser189Arg and p.Pro459Leu do not alter ZAP70 and LAT phosphorylation upon TCR stimulation of heterozygous KI Jurkats.** Cells were starved, stimulated by αCD3/αCD28 antibody crosslinking for 30 minutes, and lysed. Western blot was performed for the indicated proteins. (**A)** Representative images. (**B**-**C**) Band intensities were quantified and normalized to GAPDH or α-actinin performed on the same blots. Graphs show the ratio of normalized phospho/total ZAP70 and LAT.

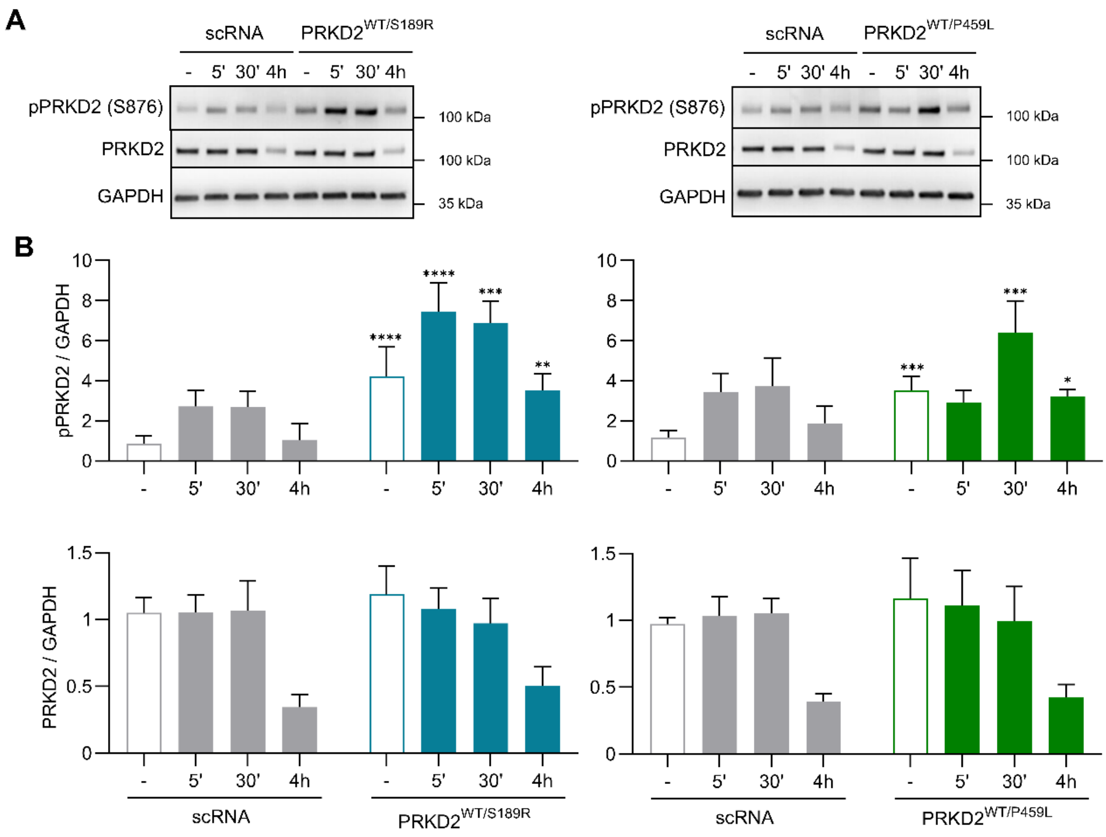


**Supplementary Figure S4: p.Ser189Arg and p.Pro459Leu show different kinetics of TCR-stimulated PRKD2 autophosphorylation in heterozygous KI Jurkats.** Cells were starved, stimulated with αCD3/αCD28 antibody crosslinking for 30 minutes, and lysed. Western blot was performed for the indicated proteins. (**A**) Representative images. (**B**) Band intensities were quantified and normalized to GAPDH performed on the same blots. Graphs show the ratio of normalized phospho- (upper) and total (lower) PRKD2.

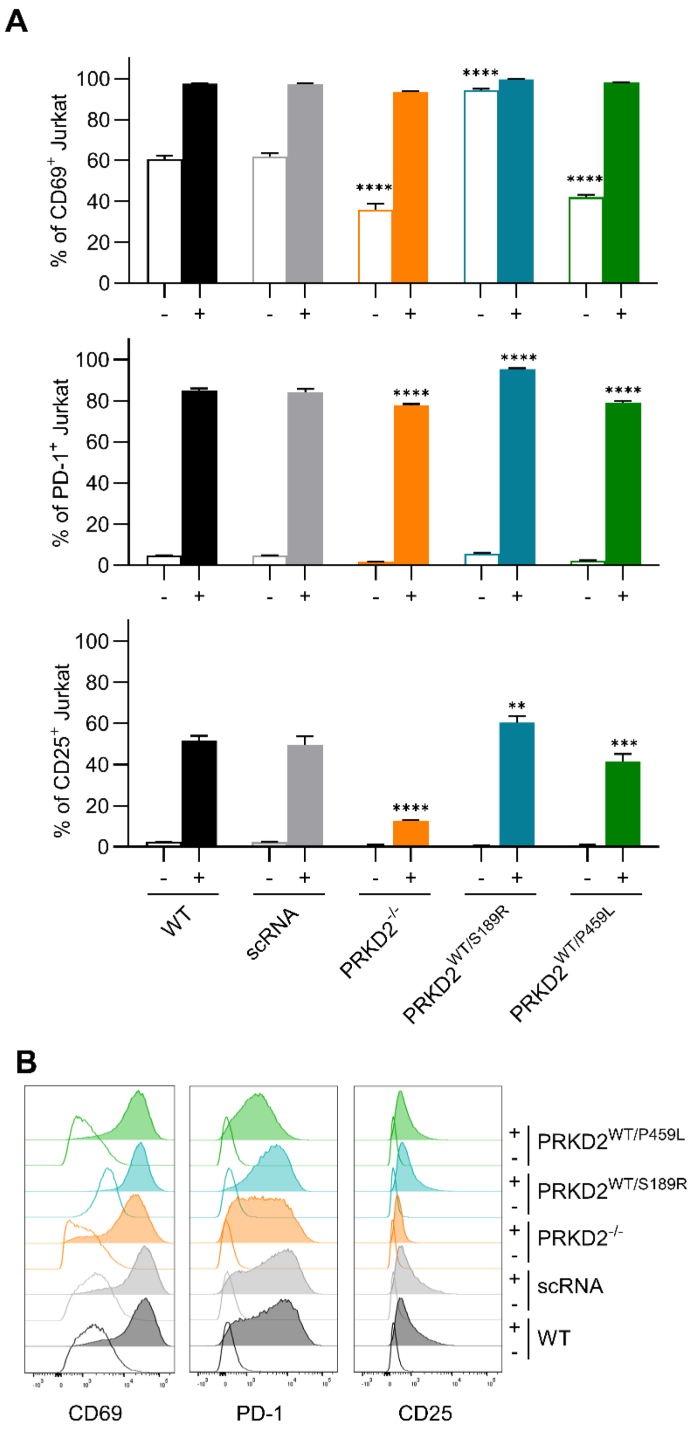


**Supplementary Figure S5: Effects of p.Ser189Arg and p.Pro459Leu on expression of T cell activation markers at 72 hours of TCR-stimulation of heterozygous KI Jurkats.** Jurkat CD4 T cell clones were serum-starved for 24hours in X-Vivo-10 media, then stimulated (+) or not (-) for 72 hours with anti-CD3 (1µg/mL), anti-CD28 (1µg/mL) and the crosslinker STAR117 (5µg/mL). (**A-B**) Flow cytometry was performed for the cell surface T cell activation markers CD69, PD1 and CD25. (**A**) Percentages and (**B**) representative histograms from n=3 experiments, each performed on 3 clones per genotype. $^{**}p<0.01$, $^{***}p<0.001$, $^{****}p<0.0001$ *vs.* corresponding WT, nested one-way ANOVA with Dunnett's post-hoc multiple comparisons test.

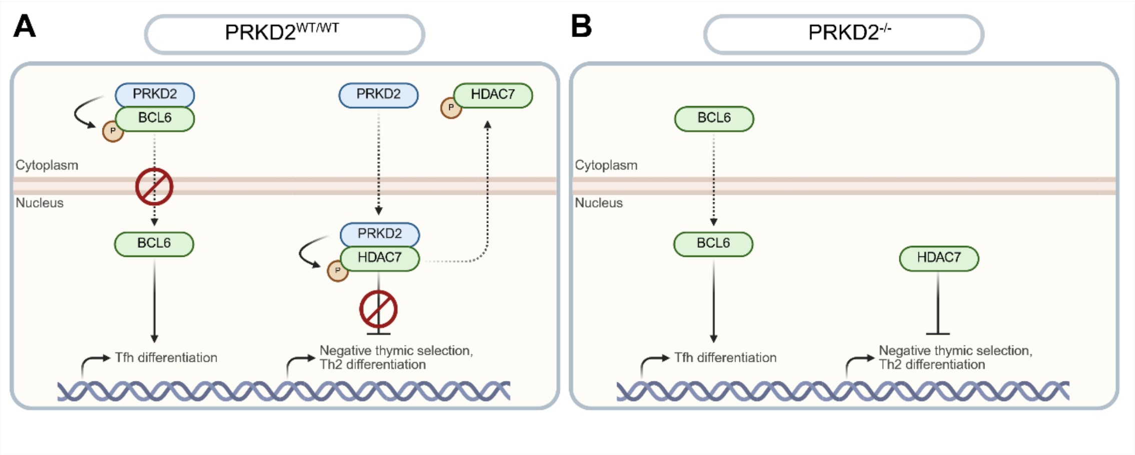


**Supplementary Figure S6: Schematic representation of the role of active PRKD2 protein in the cytoplasmic and nuclear compartments in T cells.** (**A**) In the cytoplasm, PRKD2 interacts with and phosphorylates the transcription factor BCL6, thereby limiting its nuclear translocation and the expression of genes required for $T_{FH}$ differentiation. Upon activation, PRKD2 translocates into the nucleus where it phosphorylates HDAC7, promoting its nuclear export and enabling the expression of Nur77, a key factor involved in negative selection of T cells and $T_{H2}$ differentiation. (**B**) In the absence of PRKD2, BCL6 accumulates in the nucleus, enhancing $T_{FH}$ generation, while HDAC7 is retained in the nucleus, impairing Nur77 expression and inhibiting the negative selection of T cells. Based on this, we suggest a model in which with the p.Ser189Arg variant, which increases nuclear translocation of PRKD2, permits increased nuclear entry of BCL6, promoting $T_{FH}$ differentiation. p.Pro459Leu, which decreases PRKD2 nuclear translocation, may instead affect thymic selection and promote $T_{H2}$ differentiation.
