## Supplementary Table S1 for "Activating variants in PRKD2 in two families with Systemic sclerosis, and enrichment of rare variants in a sporadic series"

| Family | Individual | Clinical description |
|---|---|---|
| A | A-1 | **A-1**, a male smoker, was diagnosed with dcSSc in his late sixties based on recent-onset Raynaud's phenomenon, scleroderma pattern on capillaroscopy, positive ANA with anti-RNA polymerase III antibodies, diffuse skin involvement, and a past episode of scleroderma renal crisis. He was treated with Rituxan with later addition of mycophenolate mofetil due to progression of skin involvement. The latter was stopped due to lymphocytic colitis, restarted for pericarditis, but discontinued shortly afterwards due to pneumonia. Disease is currently stable without evidence of interstitial lung disease and pulmonary arterial hypertension. |
| | A-2 | **A-2**, a male smoker, was diagnosed with dcSSc in his late forties based on Raynaud's phenomenon, scleroderma pattern on capillaroscopy, and diffuse skin involvement. He has a positive ANA but without SSc specific antibodies. Methotrexate was started at the moment of diagnosis with Rituxan added due to insufficient disease control. Mild fibrotic changes were observed on HRCT a few years later, and Methotrexate was stopped on the patient's request. He was re-treated with Rituxan for progression of skin involvement and synovitis, and mycophenolate mofetil was added. The patient does not show pulmonary arterial hypertension. |
| B | B-1 | **B-1**, a male smoker, was diagnosed with dcSSc in his early fifties based on Raynaud's phenomenon, scleroderma pattern on capillaroscopy, and diffuse skin involvement. At the time of diagnosis he had interstitial lung disease secondary to SSc. The patient had a positive ANA but without SSc specific antibodies. Methotrexate was started at the time of diagnosis, with Rituxan added intermittently due to progression of skin involvement. He was later diagnosed with a squamous cell lung carcinoma and is deceased. |
| | B-2 | **B-2**, a male former smoker, was diagnosed with lcSSc in his fifties, based on Raynaud's phenomenon, scleroderma pattern on capillaroscopy, anti-centromere antibodies and sclerodactyly. A repeat HRCT showed the beginnings of interstitial lung disease without restriction in lung function. He later developed ischemic cardiomyopathy without evidence for pulmonary arterial hypertension on right heart catheterization, and was hospitalized with digital necrosis. X-ray of the lungs showed pleural effusion due to a sclerosing pleuritis for which a talc pleurodesis was performed. Due to the limited and stable skin involvement no immunomodulatory medication was initiated during the course of the disease. The patient has since been lost to follow-up. |
| C | C-1 | **C-1**, a male smoker, was diagnosed in his late twenties based on Raynaud's phenomenon and scleroderma pattern on capillaroscopy. There was no skin involvement at the time, and he had a negative ANA. He later developed sclerodactyly (first non-Raynaud feature). Due to mild and stable disease over the years (lcSSc without cardiopulmonary involvement) there has been no need for immunomodulatory therapy. |
| | C-2 | **C-2**, a male smoker, was diagnosed with lcSSc in his early thirties, based on Raynaud's phenomenon, scleroderma pattern on capillaroscopy and borderline anti-topoisomerase positivity. He had mild sclerodactyly. There is no cardiopulmonary involvement. Methotrexate was started at the moment of diagnosis and later stopped. Disease has since remained stable and immunomodulatory therapy has not been employed. |
| D | D-1 | **D-1**, a female non-smoker, was diagnosed with lcSSc in her fifties based on Raynaud's phenomenon, scleroderma pattern on nailfold capillaroscopy, sclerodactyly and facial skin involvement, with positive ANA and negative ENA antibodies. Since diagnosis, disease has remained stable, with no evidence of systemic organ involvement or progression of cutaneous manifestations. To date, she has not received disease-specific therapy, apart from treatment with calcium channel blockers. |
| | D-2 | **D-2**, a female non-smoker, was diagnosed with lSSc in her teens based on Raynaud's phenomenon, scleroderma pattern on nailfold capillaroscopy, positive ANA and negative ENA antibodies. Since diagnosis, disease course has remained stable, with no progression to cutaneous or systemic involvement to date. She has not required any specific treatment. |
| E | E-1 | **E-1**, a female non-smoker, was diagnosed with lSSc in her thirties based on Raynaud's phenomenon, abnormal nailfold capillaroscopy, positive ANA, and no detectable ENA antibodies. The disease remained clinically stable for several years, after which she developed severe anemia, leading to the diagnosis of gastric antral vascular ectasia (GAVE) |

| | | |
|---|---|---|
| | | as a complication of SSc, requiring endoscopic cauterization. To date, she has not developed any cutaneous manifestations or other systemic complications. She is currently managed with iron supplementation alone. |
| | **E-2** | **E-2**, a female non-smoker, was diagnosed with lSSc in her late thirties based on Raynaud's phenomenon, telangiectasia, and upper gastrointestinal symptoms. A year later she developed distal cutaneous involvement limited to the fingers, consistent with progression to lcSSc. The disease subsequently remained stable, without additional systemic complications, and was managed with calcium channel blockers and acid-suppressive therapy. She later developed an aggressive invasive breast carcinoma and is deceased. |
| **F** | **F-1** | **F-1**, a female former smoker, was diagnosed with lcSSc in her late forties based on Raynaud's phenomenon, scleroedema, and cutaneous involvement of the fingers, hands, and forearms, with positive anti-topoisomerase I antibodies. Initial treatment included calcium channel blockers and methotrexate, with development of inflammatory arthritis and tendon friction rubs requiring escalation of methotrexate dose. Under this treatment, joint involvement has remained stable, with no additional systemic complications or progression of cutaneous manifestations to date. |
| | **F-2** | **F-2**, a female non-smoker, was diagnosed with lSSc in her early twenties based on Raynaud's phenomenon, positive anti-centromere antibodies, and abnormal nailfold capillaroscopy, without cutaneous involvement. She was treated with calcium channel blockers and acid-suppressive therapy for hearburn. A year later she developed multisystem sarcoidosis, presenting with fever, multiple lymphadenopathy, and pulmonary infiltrates, which required a 6-month course of oral glucocorticoids. At present, her condition remains stable under calcium channel blockers and acid-suppressive therapy alone, with no development of cutaneous or additional systemic complications. |

**Supplementary Table S1.** Clinical description and case histories of the affected members of six SSc families.
