## Supplementary Methods for "Activating variants in PRKD2 in two families with Systemic sclerosis, and enrichment of rare variants in a sporadic series"

### WES data primary analysis

Raw data (.fasta files) were aligned to the reference human genome assembly (GRCh38) using BWA 0.7.15[1]. Aligned sequences (.bam files) were processed using Samtools 1.12 "MarkDup" (for marking duplicates) and GATK 4.2 "BQSR" (for base quality scores recalibration). Variant calling was then performed using GATK 4.2 Haplotype Caller (following Broad Institute best practices). The variant call (.vcf) files generated were imported, annotated and further analyzed on Highlander 17.18 (https://sites.uclouvain.be/highlander/), the in-house bioinformatics framework of the Genomics and Bioinformatics platform of UCLouvain (PGEN: https://www.deduveinstitute.be/pgen-bioinformatics), with a local database currently containing more than >5800 WES.

### WES missense variant effect prediction

Missense variants were retained if ≥5 out of the following 20 tools predicted a deleterious effect: DAMAGING in Mutation Taster, FATHMM, FATHMM-XF, Polyphen2 (HDIV), Provean, SIFT4G, Mutation Assessor, MCAP, LRT, Lists2, Deogen, ClinPred, BayesDel (with MaxMAF), PrimateAl and MetaSVM; Score >20 in CADD phred, >0.5 in VEST, >0.5 in REVEL, >0.75 in MVP, >0.75 in MutPred.

### crRNA and HDR donor template sequences for generation of KO and KI Jurkat clones

| PRKD2 modification | crRNA sequence | Donor template sequence |
|---|---|---|
| KO (exon 1) | CCGATAGGAGTGCACCGTGA | None |
| p.Ser189Arg KI (exon 5) | ACTGTAGTGGGGCCCGCAAA | AGTGGCCACTGGCCAGAGACGTGGATGACAGGCGGCGTTTGCGGGCCCCTCTGCAGTTGTTGGGGATGCTGAAGGCACAGCGCTTGTGGT |
| p.Pro459Leu KI (exon 11) | GATCTCAAAGCAGTGTGGGT | AAGTAGGTGGCATTGGCAGTGACAATCTCAAAGCAGTGGGGATTGGTGCCCGGCAGCACAAGGCTGAAGTTCTGGGCGGACTCCACCGTGAGGAT |

**Red: Familial SSc variant.** **Green: Silent mutations introduced to enhance HDR by decreasing continued genome editing.**
Clones positive for p.Ser189Arg were found to also carry the accompanying silent mutations; this was not the case for p.Pro459Leu-positive clones.

**Targeted NGS-screening of KI Jurkat clones**

Genomic DNA was extracted from cells grown in 96-well plates, by boiling and digestion with 10 μg/mL Proteinase K (Merck, P2308). ~400-bp regions flanking the variant sites (in exons 5 and 11 of the *PRKD2* gene; ENSG00000105287) were amplified using KAPA HiFi DNA Polymerase (Roche, #KK2502). Primers included universal 5′ tails to enable a second PCR for incorporation of Illumina adapters and sample-specific barcodes. Amplicons were purified with 1.2X AMPure XP beads (Beckman Coulter, A63881), and indexing PCR was performed using the Nextera XT Index Kit v2 (Illumina). Following AMPure bead-purification, libraries were sequenced on an Illumina MiSeq (PGEN platform, UCLouvain) using the MiSeq Reagent Kit v2 (500 cycles; Illumina).

**Western blot**

Proteins were separated by migration of cell lysates on (4-15% gradient) polyacrylamide gels (BioRad, #4561086) in 1x Run buffer (BioRad, #1610772). Proteins were transferred onto PVDF membranes (BioRad, #162077) using 1x Transfer buffer (BioRAD, #10026938) in a semi-dry transfer system (BioRad, #27168) for 30min at 130V. Membranes were blocked in 2% milk-TBS-0.1% Tween for 1 hour at room temperature then incubated overnight at 4°C with primary antibodies diluted in 5% BSA-TBS-0.1% Tween. Membranes were washed 3x5 minutes with TBS-0.1% Tween then incubated in secondary antibody diluted in 2% milk-TBS-0.1% Tween for 1 hour at room temperature. After 3x5 minute washes with TBS-0.1% Tween and a 5 minute wash with

TBS, membranes were incubated with enhanced chemiluminescence substrate (ECL, ThermoFisher, #34580 and #34095) and images scanned on the Fusion Solo 7S Edge (Vilber).

| Primary antibody | Dilution | Company | Catalog # |
|---|---|---|---|
| GAPDH | 1/1000 | Cell Signaling | 2118 |
| α-actinin | 1/1000 | Cell Signaling | 6487 |
| pPRKD2 (Ser876) | 1/500 | Abcam | ab51251 |
| PRKD2 | 1/1000 | Cell Signaling | 8188 |
| pIκBα (Ser32) | 1/500 | Cell Signaling | 2859 |
| IκBα | 1/1000 | Cell Signaling | 4814 |
| pNF-κB p65 (Ser536) | 1/500 | Cell Signaling | 3033 |
| NF-κB p65 | 1/1000 | Cell Signaling | 4764 |
| pErk p42/44 (Thr202/Thr204) | 1/1000 | Cell Signaling | 9101 |
| Erk p42/44 | 1/1000 | Cell Signaling | 9102 |
| pAkt (Ser473) | 1/1000 | Cell Signaling | 4060 |
| Akt | 1/1000 | Cell Signaling | 9272 |
| pZAP70 (Tyr319) | 1/1000 | Cell Signaling | 2717 |
| ZAP70 | 1/1000 | Cell Signaling | 3165 |
| pLAT (Tyr220) | 1/1000 | Cell Signaling | 3584 |
| LAT | 1/1000 | Cell Signaling | 45533 |

| Secondary antibody | Dilution | Company | Catalog # |
|---|---|---|---|
| Goat anti-rabbit IgG HRP-linked | 1/10 000 | Cell Signaling | 7074 |
| Rabbit anti-mouse IgG HRP-linked | 1/10 000 | Cell Signaling | 58802 |

**Primary and secondary antibodies and dilutions used in Western blot**

### Flow cytometry

| | Antibody | Dilution | Company | Catalog # |
|---|---|---|---|---|
| **For Jurkat** | CD69-BV421 | 1/100 | BD Biosciences | 562884 |
| | PD1-BV786 | 1/100 | BD Biosciences | 329930 |
| | CD25-PE | 1/100 | BioLegend | 356104 |
| **For PBMC** | CD4-PE-Cy7 | 1/500 | BioLegend | 217414 |
| | CD19-BV605 | 1/100 | BioLegend | 302244 |
| | CD69-BV421 | 1/100 | BioLegend | 562884 |
| | PD1-BV786 | 1/100 | BD Biosciences | 329930 |
| | CD25-PE | 1/200 | BD Biosciences | 356104 |

**Fluorescence-conjugated antibodies and dilutions used in flow cytometry**

### RNASeq data analysis

Fastq files were processed using Trimmomatic (v0.39)[2] to remove low quality reads, HISAT2 (v2.2.1)[3] to align reads to the human genome (GRCh38), and evaluation of gene expression using featureCounts from Subread (v2.0.3)[4] and Homo_sapiens.GRCh38.105.chr.gtf. Differential expression analyses were performed with DESeq2 Bioconductor package v1.52.0[5]. Gene set enrichment analyses (GSEA) were performed using clusterProfiler v4.16.0[6] on genesets from the Molecular Signatures Database (MSigDB) downloaded using msigdbr package v10.0.2. RNASeq data have been deposited in the Gene Expression Omnibus (GEO, NCBI; accession number GSE346025).
